# An AI-Integrated Framework for Precision Genomics in Coronary Artery Disease Using Whole Exome and Phenotypic Data

**DOI:** 10.64898/2026.01.28.26345099

**Authors:** Kalyan Ram Uppaluri, Hima Jyothi Challa, Krishna Kumar Vempati, Lakshmi Nirmala Kadali, Madhubhanu Rayala, Kalyani Palasamudram

## Abstract

Coronary artery disease (CAD) is a multifactorial condition influenced by genetic, phenotypic, and environmental factors. Traditional risk prediction models fall short in capturing the polygenic complexity of CAD, particularly in underrepresented populations. This study presents SIGMA (Scoring Importance of Genes specific to disease using Machine learning Algorithms), a novel AI-powered framework that enhances CAD risk prediction by integrating genomic and phenotypic data. Our approach leverages GEMS (GeneConnectRx Evidence Metrics), an LLM-driven system to score 1772 CAD-associated genes, and CASCADE (Comprehensive Assessment of Sequence and Clinical Annotation Data Evaluation), a tiered variant scoring pipeline. Using whole exome sequencing (WES) data from 1,243 individuals (628 controls, 615 CAD cases), the model integrates age and gender as key non-modifiable phenotypes. Results show significant improvements in sensitivity (from 0.41 to 0.79), specificity (0.70 to 0.72), and AUC (0.59 to 0.81) when phenotype data are incorporated. Our findings highlight the potential of AI-integrated genomics for population-specific CAD risk stratification.

## 1. Introduction

CAD remains a leading global cause of cardiovascular morbidity and mortality, presenting complex challenges in risk stratification and early intervention. While traditional CAD prediction models primarily rely on phenotypic factors and biochemical parameters, incorporating genetic factors has always been challenging because of the polygenic nature of the disease (1). The main challenges of incorporating genetic data include multiple genes being involved in the disease pathogenicity, variants in each gene having a cumulative effect (2), missing heritability (3), the impact of each variant on the disease, the importance of common vs rare variants on phenotype (4), and epigenetic mechanisms on gene expression (5) to name a few.

To address these complexities, we developed a novel integrative framework—**SIGMA**—that leverages Machine learning and LLMs to enhance CAD risk prediction by incorporating genetic and phenotypic inputs.

[Figure 1] shows the pipeline with biological context and literature-driven gene identification, followed by systematic variant annotation, prioritization, and scoring. We introduce **GEMS**, an LLM-based tool for evaluating gene-disease associations, and **CASCADE**, a multi-tiered genomic variant scoring framework. These tools enable personalized CAD risk estimation, combining literature-informed gene relevance, ACMG-guided variant classification, and population-specific data interpretation.

**Figure 1:**
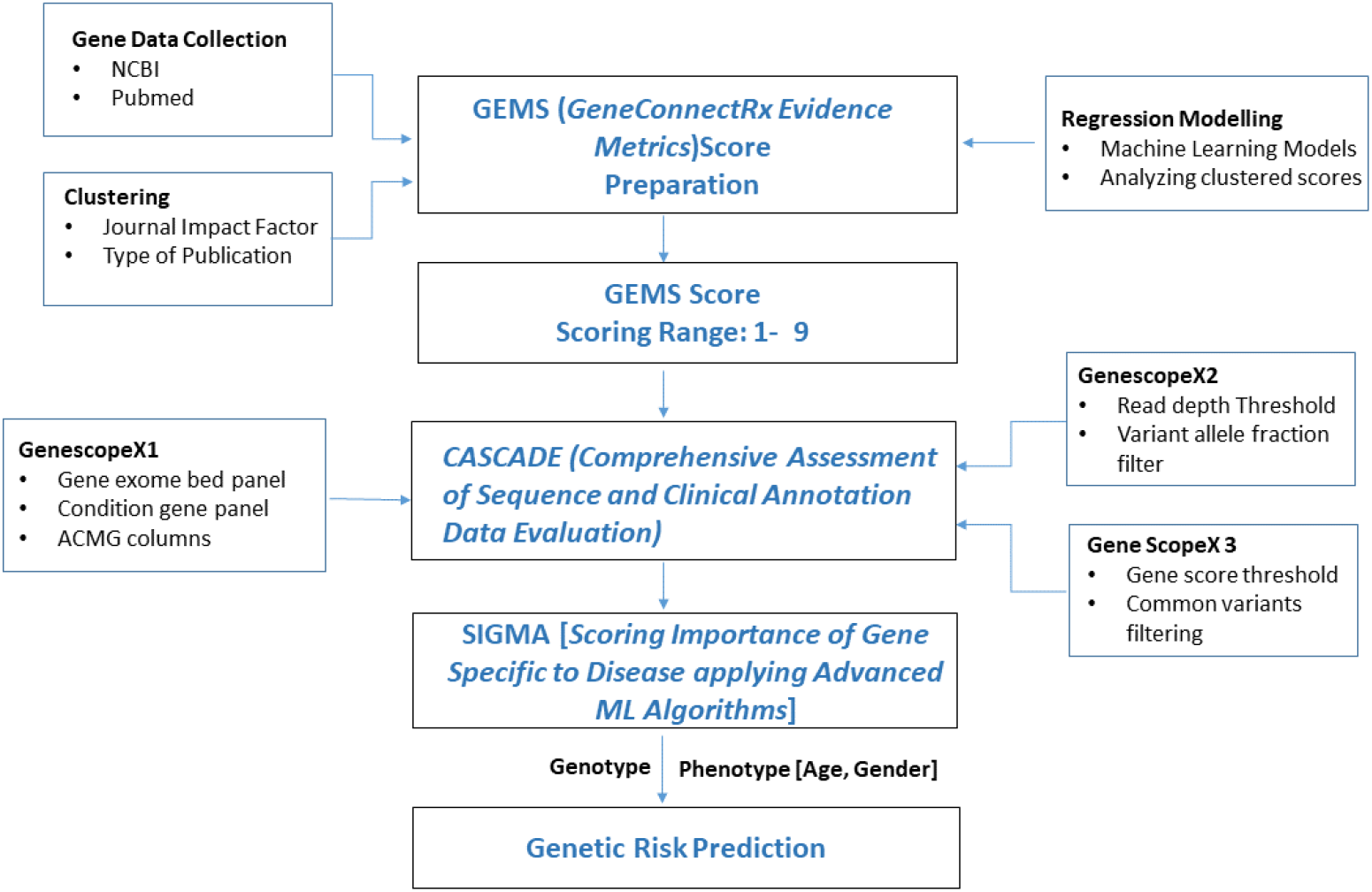
End-to-End Pipeline: From data collection to Genetic Risk Prediction. This figure outlines the end-to-end workflow for predicting the genetic risk of CAD. It begins with gene data collection from sources like NCBI and PubMed, followed by scoring through the GEMS system. The GEMS score is refined using clustering, regression models, and filters from various gene panels (GenoscopeX1, X2, X3). These scores are evaluated through CASCADE and SIGMA modules. Finally, combining genotype data with phenotype factors (age, gender) enables personalized genetic risk prediction.

## 2. Methodology

The primary objective is to enhance model performance by evaluating sensitivity and specificity by integrating genetic and phenotypic data. Genetic data helps identify inherent risks, while phenotypic data encompasses personal non-modifiable factors such as age and gender.

### 2.1 Study Population and Data Collection

A total of 1,243 subjects were enrolled through GenepoweRx under informed consent. WES was conducted for all participants. The cohort included:

- Cases (n=615): Individuals who underwent PCI with either balloon dilatation and/or stent placement or coronary artery bypass procedure.
- Controls (n=628): Age- and gender-matched individuals with normal angiograms and/or stress tests.

Demographic data and non-modifiable Phenotype factors (age, gender) were recorded for all participants.

### 2.2 GEMS: Literature-Driven Gene Prioritization via Large Language Models

GEMS (GeneConnectRx Evidence Metrics) is a novel AI-powered platform designed to systematically quantify the strength of gene-disease associations using published scientific literature. Recognizing that coronary artery disease (CAD) is influenced by a wide array of genetic factors, GEMS addresses the challenge of prioritizing relevant genes from vast and heterogeneous biomedical publications.

At its core, GEMS harnesses a large language model (LLM) to extract, interpret, and contextualize biomedical knowledge from peer-reviewed literature. Unlike conventional keyword-based search tools, GEMS applies natural language understanding to capture nuanced relationships between genes and disease phenotypes, including indirect or inferred links. This enables a more comprehensive and biologically relevant assessment of gene-disease relevance.

The platform incorporates a multi-layered analytical framework that evaluates not just the presence of gene mentions but the quality, credibility, and specificity of supporting evidence. This is operationalized through the following workflow (illustrated in [Figure 2]):

**Figure 2:**
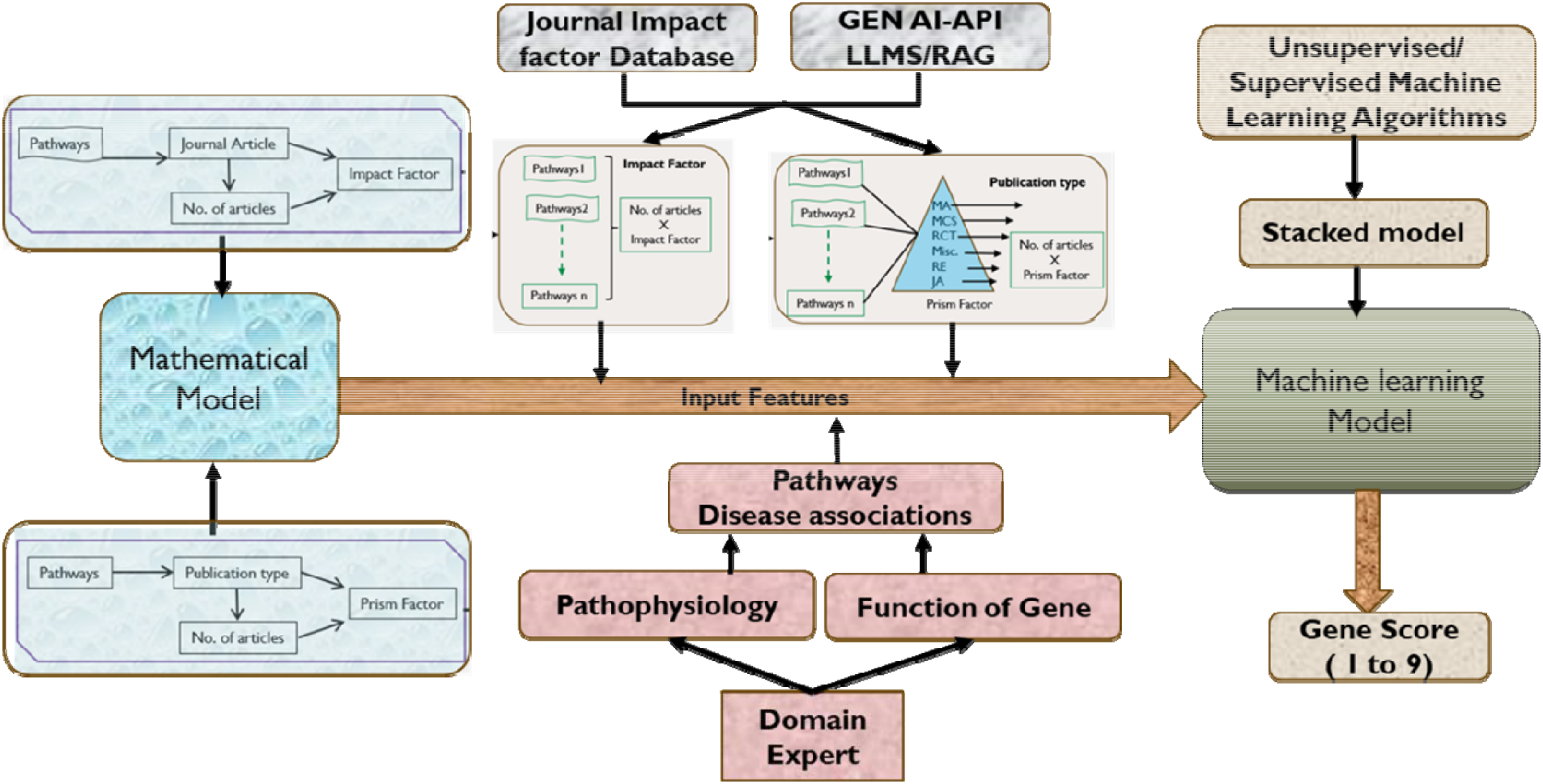
GEMS (GeneConnectRx Evidence Metrics) Framework. This figure represents the GEMS framework for gene scoring. It integrates data from journal impact factors, APIs, and expert inputs to extract features such as pathway relevance, publication count, and article types. These features are fed into machine learning algorithms (supervised/unsupervised) to generate a gene score ranging from 1 to 9, reflecting the strength of gene-disease associations.

- Data Collection: Scientific articles and genomic annotations are harvested from authoritative biomedical databases such as PubMed via the NCBI API. The LLM is employed to extract semantic associations between genes and CAD, including modifiers like epigenetic influence, comorbidities (e.g., diabetes), and disease subtypes.
- Clustering and Relevance Filtering: Extracted literature is clustered using unsupervised learning techniques that factor in journal impact metrics, publication type (e.g., meta-analysis vs. case study), citation count, and semantic proximity to CAD-related terms. This ensures that highly cited, high-impact, and biologically relevant sources are prioritized in downstream scoring.
- Regression-Based Scoring Model: The clustered literature is passed through a supervised regression model trained to predict the evidentiary strength of gene-disease associations. The model integrates multiple features—including frequency of co-mention, strength of association verbs, and domain-specific context—to assign a weighted relevance score.
- GEMS Score Assignment: Each of the 1772 genes curated from this process is assigned a final GEMS Score ranging from 1 to 9. A score of 9 indicates high-confidence, literature-rich associations with CAD pathophysiology; a score of 1 represents limited or weak evidence. The gene set includes those implicated in CAD directly, in CAD-equivalent conditions such as diabetic macrovascular complications, and in upstream epigenetic regulation of cardiovascular traits.

This structured scoring system allows for transparent, data-driven gene prioritization, enabling downstream variant annotation pipelines (such as CASCADE) to operate with higher specificity and biological relevance. By automating the labor-intensive process of literature review through LLMs, GEMS introduces a scalable and reproducible approach to gene selection for complex disease modeling. We agree that large language models (LLMs) can generate variable outputs when not appropriately constrained. To avoid this, our gene-evidence scoring and variant-prioritization pipelines do not rely on generative, unconstrained LLM outputs. Instead, both GEMS and CASCADE use rule-based frameworks, curated databases, and deterministic evidence-aggregation methods.

Specifically: GEMS does not generate associations; it aggregates them. The LLM component is restricted to summarizing already-established evidence, not creating new gene–disease links. No novel claims depend on LLM inference. All reported gene–CAD associations are derived from peer-reviewed sources. The LLM is used only to organize and summarize evidence already present in structured datasets. All high-confidence GEMS associations and top-tier CASCADE variants were cross-checked manually, ensuring consistency across runs.

Thus, the workflow avoids the primary risks associated with stochastic LLM behavior. The system functions as a controlled evidence-synthesis tool, not a generative inference engine.

### 2.3 CASCADE: A Robust Variant Scoring Framework for Ethnicity-Specific CAD Risk Prediction

CASCADE (Comprehensive Assessment of Sequence and Clinical Annotation Data Evaluation) is a three-step variant filtering and scoring pipeline designed to generate an ethnicity-specific, biologically meaningful genetic risk score for coronary artery disease (CAD). It prioritizes genetic variants using curated gene associations, clinical impact annotations, and population-specific allele frequencies. This framework is especially suited to South Asian populations, addressing a critical gap in the generalizability of existing polygenic risk models developed in European cohorts.

[Figure 3] presents an overview of the CASCADE pipeline, which operates on annotated variant data from whole exome sequencing (WES) and integrates gene relevance from the GEMS platform.

**Figure 3:**
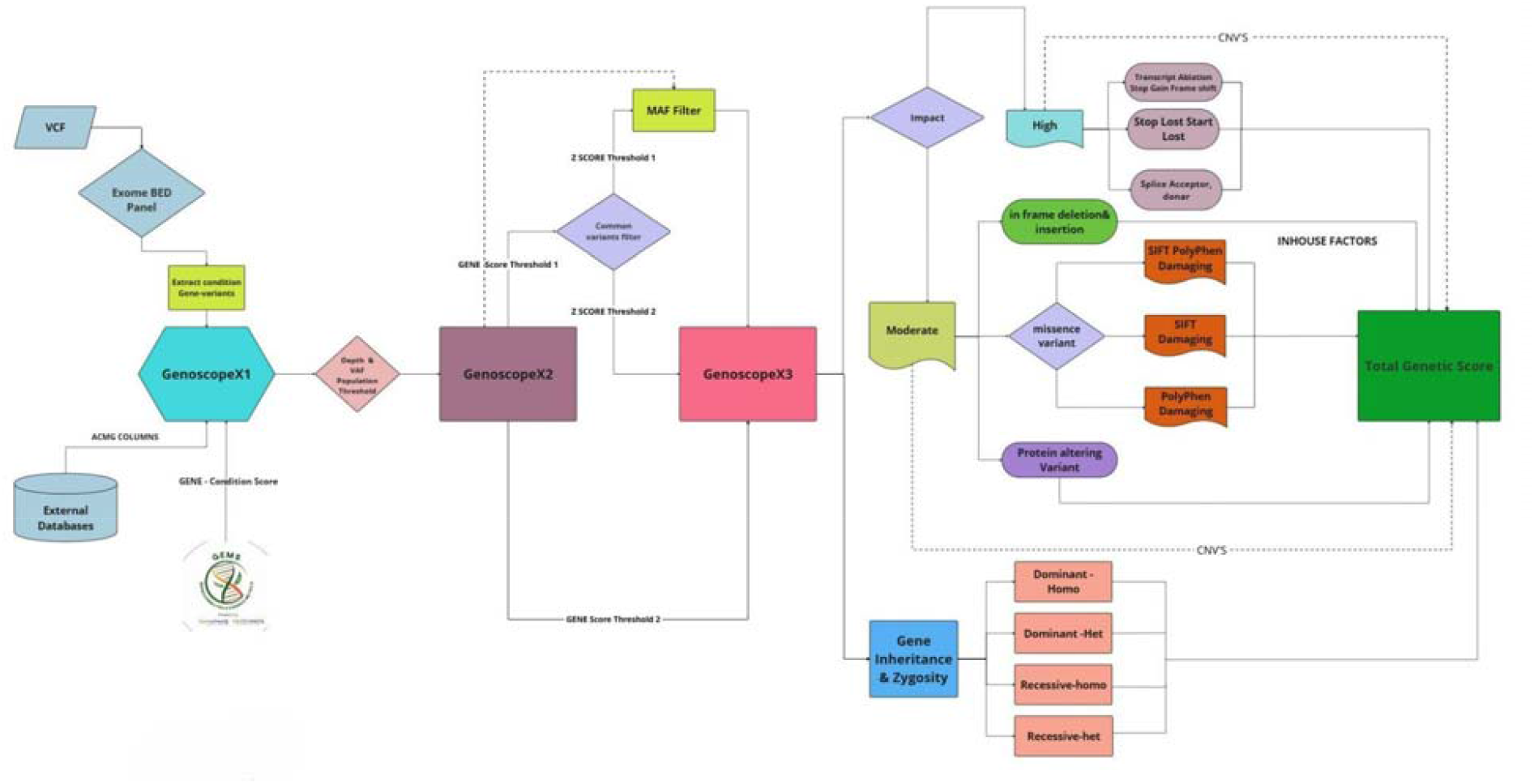
Overview of the CASCADE Workflow. This figure outlines the CASCADE workflow—a stepwise approach to genomic scoring. It begins with variant data (VCF), processed through GenoscopeX1, X2, and X3 modules that apply quality filters like gene panel mapping, read depth, allele frequency, and score thresholds. It further integrates variant impact, inheritance patterns, zygosity, and phenotypic relevance to compute the Total Genetic Score

#### Step 1: Variant Extraction and Functional Annotation (GenescopeX1)

Raw FASTQ files were assessed using FastQC and MultiQC. After alignment (BWA-MEM), GATK Picard was used for marking duplicates, base recalibration, and indel realignment according to GATK best practices.

Sample-level QC metrics:

- Total reads, percent mapped reads, on-target rate
- Mean depth and uniformity of coverage (≥80% of bases within 0.2× of the mean)
- Transition/transversion ratio (Ti/Tv) expected range 2.6–3.1
- Heterozygosity rate outlier removal (±3 SD)
- VerifyBamID2 contamination estimate <3%
- Sample identity confirmation (genotype fingerprinting across markers)

Variant-level QC metrics:

- Hard filters for low-quality SNVs/indels
- VQSR tranche sensitivity 99.0% threshold
- Depth, genotype quality, strand bias, MQRankSum, ReadPosRankSum thresholds
- Removal of multi-allelic sites if any allele failed QC
- PASS-only variants accepted

Common sequencing artefacts (oxidative damage, homopolymer tails) were removed using machine-learning filters incorporated in GATK.

The initial stage of CASCADE involves extracting variants from VCF files generated by the Variant Effect Predictor (VEP), using the Hyper KAPA Roche BED design panel.

To ensure biological relevance:

- **Only genes with strong or moderate evidence from GEMS (Gene Score** ≥ **threshold)** are considered.
- Annotations are enriched using **ACMG (American College of Medical Genetics and Genomics) guidelines**, classifying variants as either **HIGH IMPACT** (e.g., frameshift, splice-site, nonsense) or **MODERATE IMPACT** (primarily missense).

For MODERATE IMPACT missense variants, **SIFT** and **PolyPhen** predictions are incorporated to assess the likelihood of damaging effects on protein function.

Each variant record in GenescopeX1 includes:

- Genomic attributes (CHROM, POS, REF, ALT, rsID)
- Functional annotations (GeneInfo, ClinVar significance, ACMG impact)
- Prediction scores (SIFT, PolyPhen)
- Disease-specific relevance (CAD tagging based on curated gene panels)

This step ensures that only functionally relevant and literature-supported variants are passed downstream.

#### Step 2: Technical Quality Filtering (GenescopeX2)

The second stage enhances **variant reliability** by applying stringent quality control filters:

- **Depth of sequencing (DP)** ensures adequate read support.
- **Variant Allele Frequency (VAF)** thresholds remove low-confidence calls.

This generates a refined dataset—**GenescopeX2**—comprising technically high-confidence variants suitable for modeling and clinical interpretation.

#### Step 3: Rare Variant Enrichment and Weighted Scoring (GenescopeX3)

The final step refines the data using population-specific and clinical impact metrics:

- **MAFs (Minor Allele Frequencies)** are derived directly from the study cohort of 1,243 Indian individuals—addressing a major limitation of global databases, which often lack adequate South Asian representation.
- **Z-score comparisons** are used to evaluate allele frequency shifts in CAD vs. control groups.
- **Copy number variations (CNVs)** near CAD-relevant genes are also assessed and scored based on their genomic location and size.

The **Total Genetic Risk Score** is calculated by integrating:

- **Gene relevance score** from GEMS
- **Variant impact** (HIGH vs. MODERATE)
- **Pathogenicity predictions** (SIFT, PolyPhen)
- **Zygosity status** (heterozygous vs. homozygous)
- **Inheritance patterns** (dominant vs. recessive)

Variants with **HIGH IMPACT** (e.g., nonsense, splice-site, CNVs) are given the most weight. MODERATE IMPACT variants, especially damaging missense variants, are evaluated using predictive tools and included when relevant. This nuanced consideration of functional consequence, zygosity, and gene-specific inheritance modes ensures that CASCADE does not over- or under-estimate risk from any one type of variant.

Although CASCADE uses standardized gene-level and variant-level annotations (ACMG impact, GEMS gene scores, SIFT/PolyPhen predictions), the final score is not static across individuals. CASCADE integrates each participant’s unique constellation of variants, including zygosity, variant consequence, population-specific MAF shifts, and gene-level weighting, to compute a personalized Total Genetic Risk Score (TGRS). This results in a distinct quantitative score for every subject, reflecting individual genomic burden. These per-individual scores, not the underlying static annotations, are used as features for model training. Therefore, CASCADE produces individualized genomic predictors that vary across cases and controls, supporting meaningful disease classification.

### 2.4 2Population Characteristics and Risk Stratification Variables

The distribution of CAD status in the studied Indian population revealed an almost balanced cohort, comprising 1,243 individuals:

- CAD Controls: 628 individuals (50.2%)
- CAD Cases: 615 individuals (49.8%)

This near-equitable split provides an ideal foundation for training machine learning (ML) models, enabling robust detection of genomic and phenotypic features associated with disease risk [Figure 4a].

**Figure 4:**
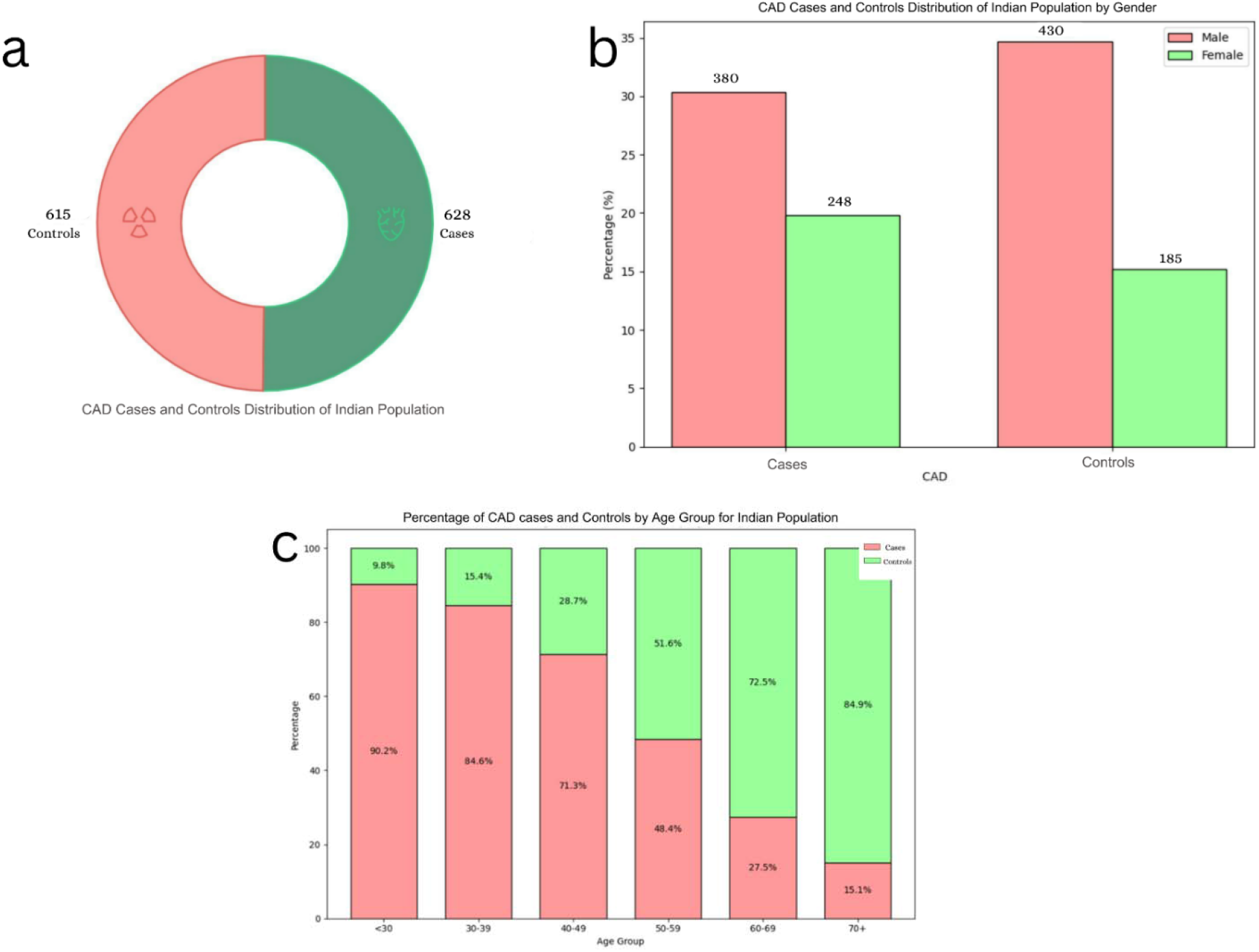
Demographic Distribution of Coronary Artery Disease (CAD) Cases and Controls in the studied Indian Population. A. Donut chart showing overall distribution of participants: 628 CAD cases and 615 controls, indicating a nearly balanced dataset. B. Bar chart representing gender-wise distribution of CAD cases and controls. CAD cases included 380 males and 248 females, while controls comprised 430 males and 185 females. The prevalence of CAD was higher among males. C. Stacked bar chart depicting the age-wise percentage distribution of CAD cases and controls. The proportion of CAD cases was higher in younger age groups (<50 years), while controls dominated in older age groups (60+ years), suggesting an age-associated risk distribution pattern.

Gender-Stratified Analysis

A higher prevalence of CAD was observed in males compared to females:

- Males with CAD: 430 (34.7% of total participants)
- Females with CAD: 185 (15.2% of total participants)

Statistical analysis revealed that males were 1.52 times more likely to develop CAD than females (Odds Ratio = 1.52, p = 0.001). Conversely, females exhibited a protective effect, with an OR of 0.56 (p = 0.001) [Table 1, Figure 4b].

**Table 1:** Comparison of Age and Gender Differences Between CAD Cases and Controls.

| Parameter | Cases (n=634) |  | Controls(n=615) |  |  |  |
| --- | --- | --- | --- | --- | --- | --- |
|  | Age(Mean ±SD) | Age(Min-Max) | Age(Mean ±SD) | Age(Min-Max) | T-test | Odds Ratio (p-value) |
| Male (m=810) | 59.3 ± 10.4 | 26-85 | 47.5 ± 11.3 | 20-83 | p=e-37 | 1.52 (p=0.001) |
| Female (m=433) | 58.2 ± 10.3 | 27-80 | 48.8 ± 11.1 | 19-75 | p=e-37 | 0.56 (p=0.001) |

Age-Stratified Analysis

Age emerged as a strong and statistically significant risk factor.

- Among males, CAD cases had a mean age of 59.3 ± 10.4 years, while controls averaged 47.5 ± 11.3 years
- Among females, CAD cases averaged 58.2 ± 10.3 years, versus 48.8 ± 11.1 years in controls The differences in both subgroups were highly significant (p ≈ 1e-37), based on independent t-tests [Table 1].

Further bin-wise analysis showed a clear trend: the likelihood of CAD increased progressively with age, reinforcing age as a primary risk determinant. These findings align with established cardiovascular epidemiology literature [Figure 4c].

### 2.5 Progressive Development of CAD Prediction Models

The model development followed a stepwise integration strategy, beginning with genetic data and subsequently incorporating phenotype information for improved performance.

Stage 1: Genotype-Only Model

The initial model utilized Total Genetic Scores computed from the CASCADE pipeline, which evaluated 1772 genes scored by GEMS. Variants were scored based on functional impact, frequency, zygosity, and inheritance, as described in earlier sections.

Stage 2: Genotype + Phenotype Integration

To enhance predictive capacity, we augmented the dataset with two non-modifiable phenotype features:

- Age
- Gender

These were selected for their well-documented clinical relevance, particularly in South Asian populations. The features were standardized using StandardScaler to ensure compatibility across ML algorithms.

Modeling Strategy

- A consistent 70:30 train-test split was used to evaluate model performance.
- Both linear (e.g., logistic regression) and non-linear classifiers (e.g., random forests, gradient boosting) were explored.
- A stacked ensemble model was constructed by integrating the best-performing classifiers from prior stages. This ensemble architecture allowed the model to leverage strengths from multiple algorithms, reducing the risk of overfitting or bias.

Performance Metrics and Model Optimization

Key performance indicators—including F1 Score, sensitivity, specificity, and AUC-ROC—were used to fine-tune model hyperparameters. The metrics were carefully monitored to ensure the final model delivered:

- Balanced class prediction
- High discriminative power
- Generalizable performance across validation folds

Two model configurations were ultimately compared (as shown in [Figure 5]):

**Figure 5:**
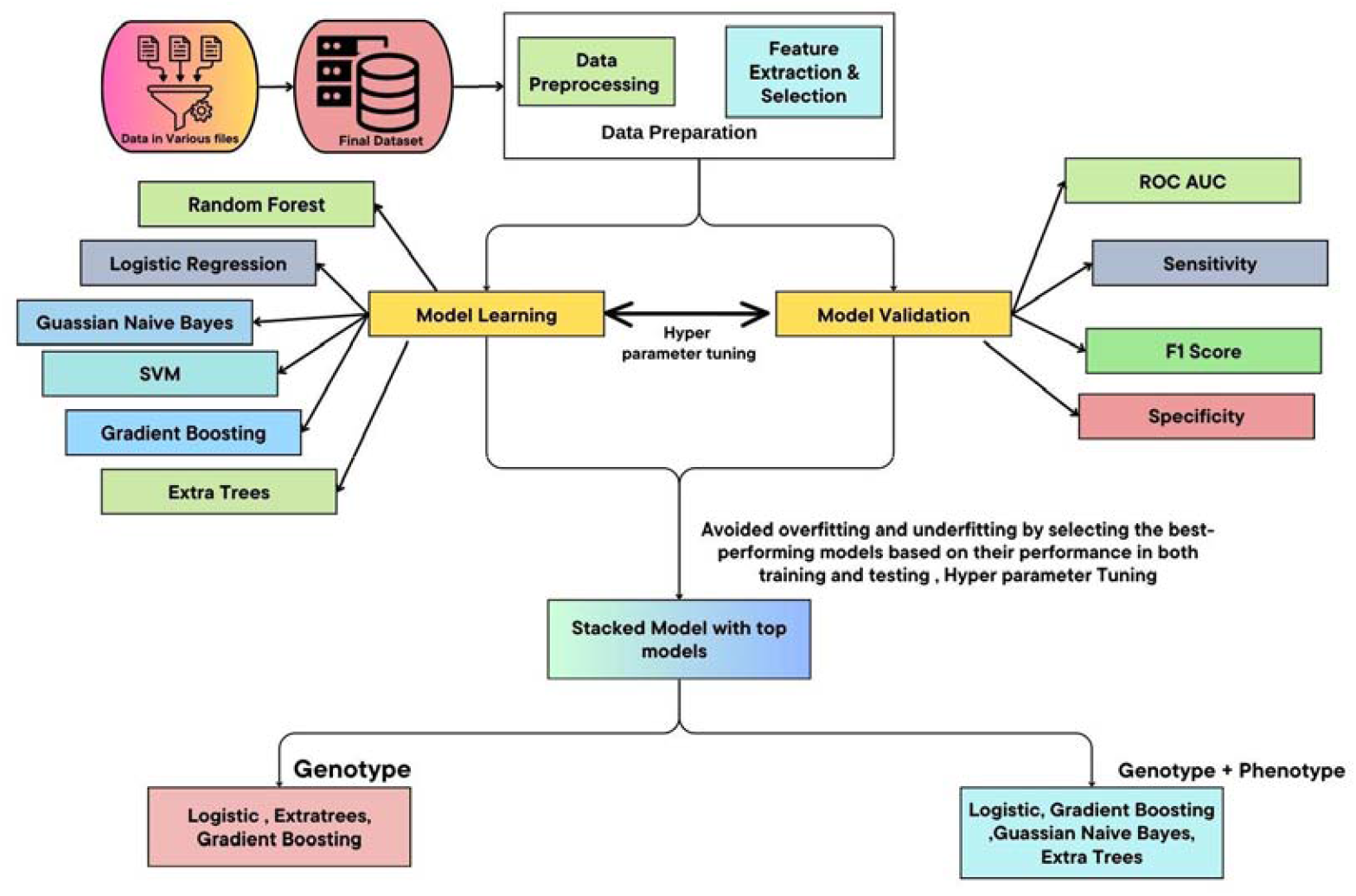
Comprehensive Machine Learning Pipeline for CAD Risk Prediction. This flowchart depicts the full workflow from raw data integration and preprocessing, which was obtained from CASCADE to model training, validation, and final classification. It highlights model selection, hyperparameter tuning, and performance metrics for genotype-only and combined genotype-phenotype predictions using ensemble and stacked models.

1. Genotype-only
2. Genotype + Phenotype (Age, Gender)

This progressive modeling approach demonstrated the additive predictive value of phenotypic variables over genomic data alone, reinforcing the importance of integrated models in personalized CAD risk assessment.

### Results

#### Model Evaluation and Performance Metrics

To systematically evaluate model performance, we used four primary metrics:

- **F1 Score**: Measures the balance between precision and recall
- **Sensitivity (Recall)**: Measures the ability to correctly identify true positives (CAD cases)
- **Specificity**: Measures the ability to correctly identify true negatives (controls)
- **AUC-ROC**: Quantifies overall discriminative performance across classification thresholds

We compared two configurations of the stacked machine learning model **(Figure 6)**.

**Figure 6:**
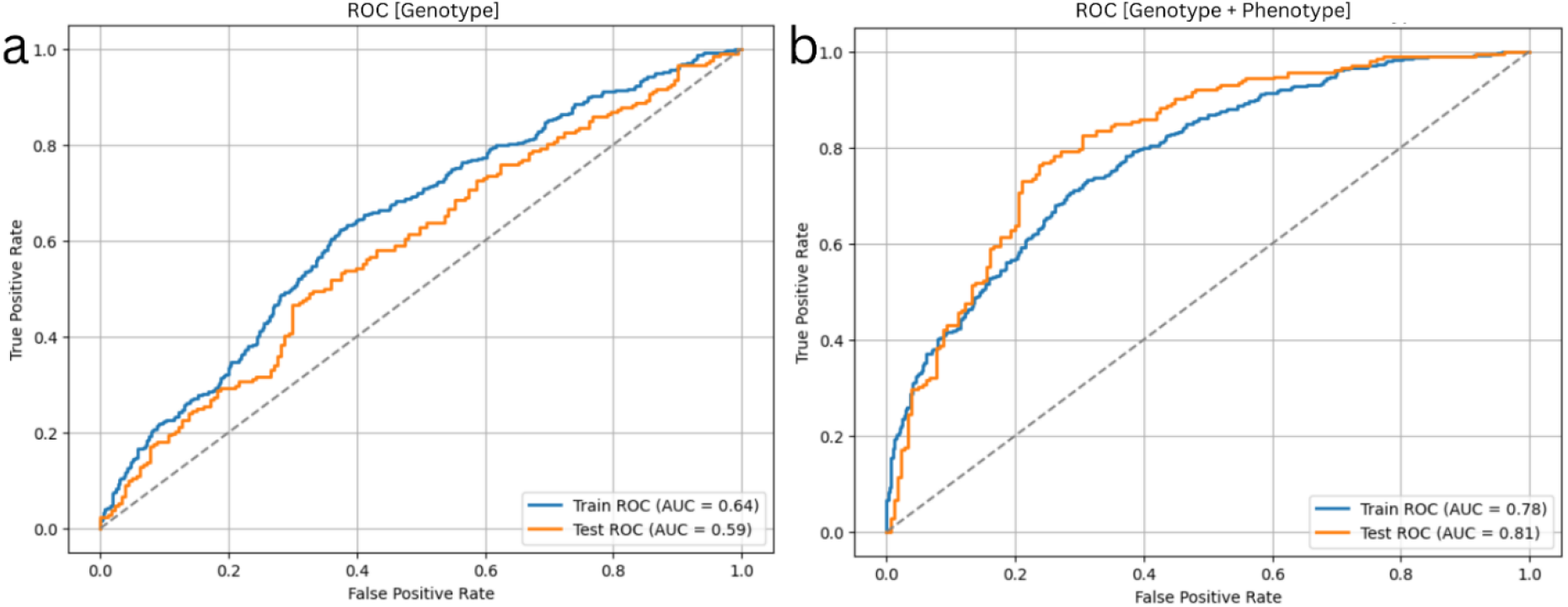
Comparative ROC Curves for Stacked Machine Learning Models Using Different Data Configurations. (a) ROC curve using only genotype data shows moderate predictive performance (Train AUC = 0.64, Test AUC = 0.59). (b) Incorporating both genotype and phenotype data (age, gender) significantly improves prediction accuracy (Train AUC = 0.78, Test AUC = 0.81), highlighting the value of the combination of Genotype + Phenotype

**Figure 7:**
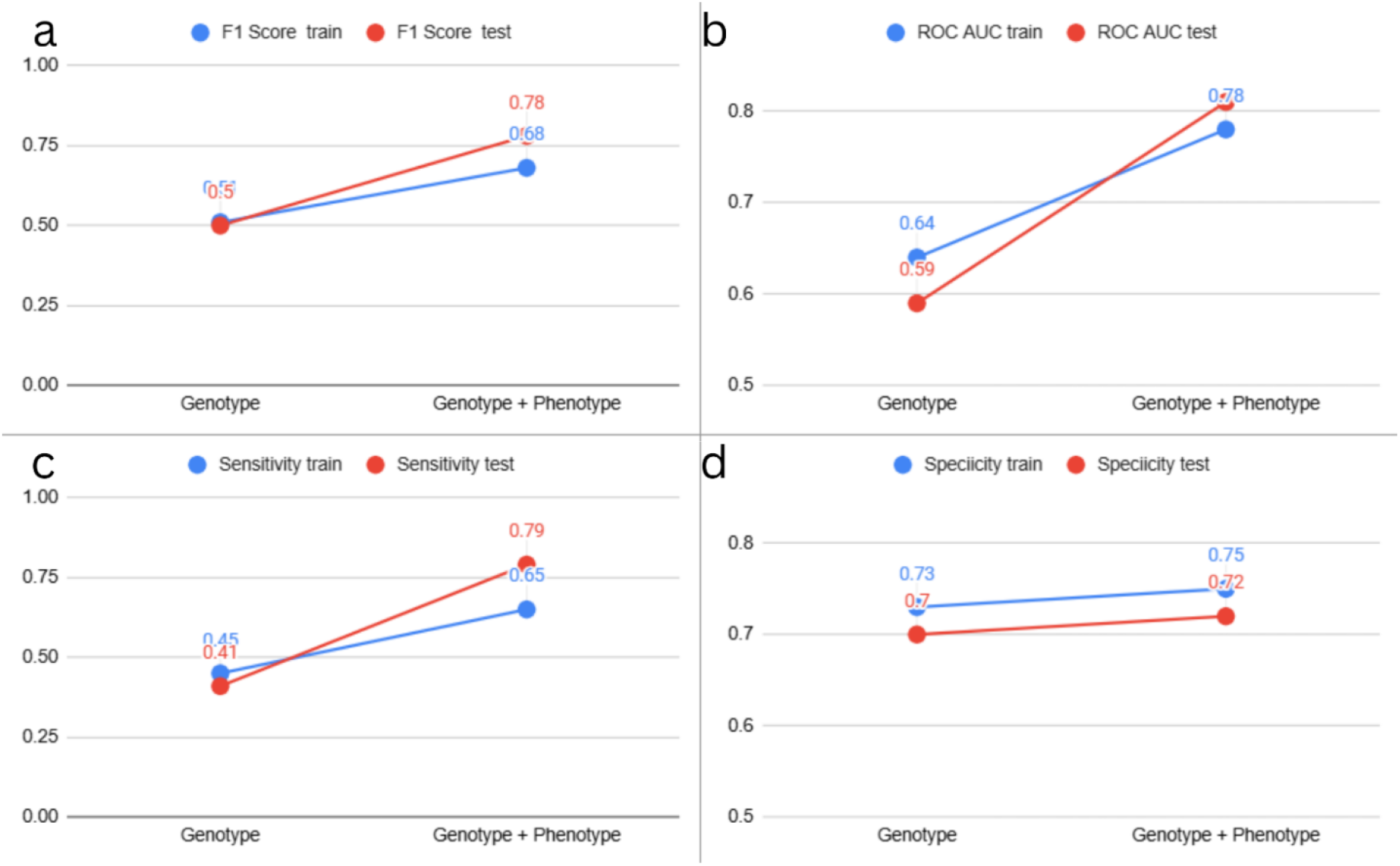
Comparative Performance Metrics for Stacked ML Models Under Different Data Configurations. a. F1 Score for training and test sets using three configurations: (i) Genotype (CAD only), (ii) Genotype (CAD + Risk Factors), and (iii) Genotype + Phenotype. b. ROC AUC for training and test sets under the same three configurations, c. Sensitivity comparison, showing how well each model correctly identifies positive CAD cases across the three data setups. d. Specificity comparison, indicates how accurately each model distinguishes non-CAD cases.

1. **Genotype-only** model using CASCADE-derived genetic scores
2. **Genotype + Phenotype** model incorporating non-modifiable features (age and gender)

**Table 2** presents a side-by-side comparison of performance metrics on the training and test datasets.

**Table 2:** Comparative Performance Metrics for Genotype VS Genotype + Phenotype Integrated Models.

| Metric | Data Configuration | Training Set | Test Set |
| --- | --- | --- | --- |
| F1 Score | Genotype | 0.51 | 0.5 |
|  | Genotype + Phenotype | 0.68 | 0.78 |
| Sensitivity | Genotype | 0.45 | 0.41 |
|  | Genotype + Phenotype | 0.65 | 0.79 |
| Specificity | Genotype | 0.73 | 0.70 |
|  | Genotype + Phenotype | 0.75 | 0.72 |
| ROC AUC | Genotype | 0.64 | 0.59 |
|  | Genotype + Phenotype | 0.78 | 0.81 |

#### F1 Score: Improved Balance Between Precision and Recall

The F1 Score, a harmonic mean of precision and recall, was chosen as the central evaluation metric due to its robustness in handling class imbalance and its balanced treatment of false positives and false negatives.

- **Training Set**: Improved from **0.51 (Genotype-only)** to **0.68 (Integrated model)** — a 35% relative gain.
- **Test Set**: Improved from **0.50 to 0.78** — a 56% increase, demonstrating strong generalization.

#### AUC-ROC: Enhanced Discriminative Power

The Area Under the Receiver Operating Characteristic Curve (AUC-ROC) assesses the model’s ability to separate CAD cases from controls across all thresholds.

- **Training Set**: AUC rose from **0.64 to 0.78** — a 22% improvement.
- **Test Set**: Increased significantly from **0.59 to 0.81**, showing a 37% relative gain in predictive discrimination.

**Sensitivity: Better Detection of CAD Cases**

Sensitivity (recall) evaluates how effectively the model identifies true positives. This is especially critical in clinical settings to reduce missed diagnoses.

- **Training Set**: Increased from **0.45 to 0.65** (44% improvement)
- **Test Set**: Jumped from **0.41 to 0.79**, a remarkable **93% increase**, highlighting the value of phenotype integration.

**Specificity: Stable True Negative Classification**

Specificity improved modestly but consistently, reflecting the model’s ability to control false positives while boosting sensitivity.

- **Training Set**: Rose from **0.73 to 0.75**
- **Test Set**: Increased from **0.70 to 0.72**

Although these improvements were smaller in magnitude, they indicate that the model preserved its ability to accurately identify control samples while expanding true positive detection.

#### Interpretation and Insights

These results underscore two key findings:

**Genetic data alone** provides a strong, biologically grounded foundation for CAD risk modeling.

- **Integrating minimal phenotypic data (age and gender)** substantially boosts performance across all metrics, demonstrating that even basic clinical information enhances the interpretive power of genomic data.
- The integrated model exhibited clearer **class separation**, reduced **prediction overlap**, and significantly improved **recall without sacrificing specificity**, making it both **clinically relevant** and **statistically robust**.

A detailed summary of the final model’s performance with the data configuration for the integrated model, including mean values, CIs, and approximate SDs (derived from the CI width) for train and test metrics, is provided in **Table 3 and Table 4:**

**Table 3:**
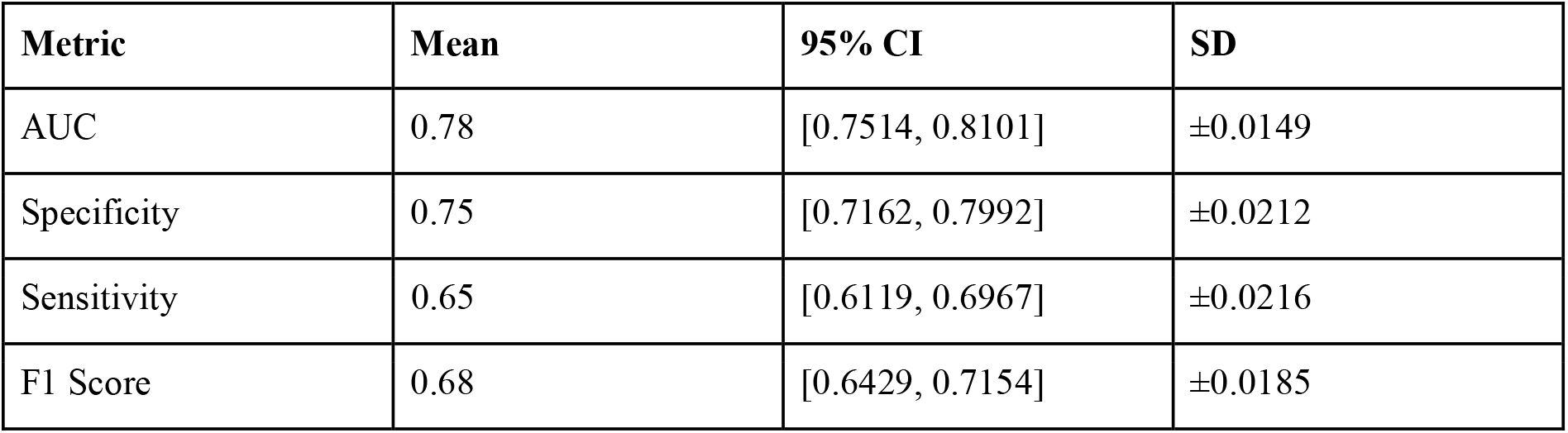
Performance metrics on the training dataset averaged over 200 bootstrap iterations.

| Metric | Mean | 95% CI | SD |
| --- | --- | --- | --- |
| AUC | 0.78 | [0.7514, 0.8101] | ±0.0149 |
| Specificity | 0.75 | [0.7162, 0.7992] | ±0.0212 |
| Sensitivity | 0.65 | [0.6119, 0.6967] | ±0.0216 |
| F1 Score | 0.68 | [0.6429, 0.7154] | ±0.0185 |

**Table 4:** Performance metrics on the test dataset averaged over 200 bootstrap iterations.

| Metric | Mean | 95% CI | SD |
| --- | --- | --- | --- |
| AUC | 0.81 | [0.7672, 0.8558] | ±0.0227 |
| Specificity | 0.72 | [0.6647, 0.7879] | ±0.0313 |
| Sensitivity | 0.79 | [0.7313, 0.8455] | ±0.0292 |
| F1 Score | 0.78 | [0.7373, 0.8235] | $\pm 0.0221$ |

**Table 5:**
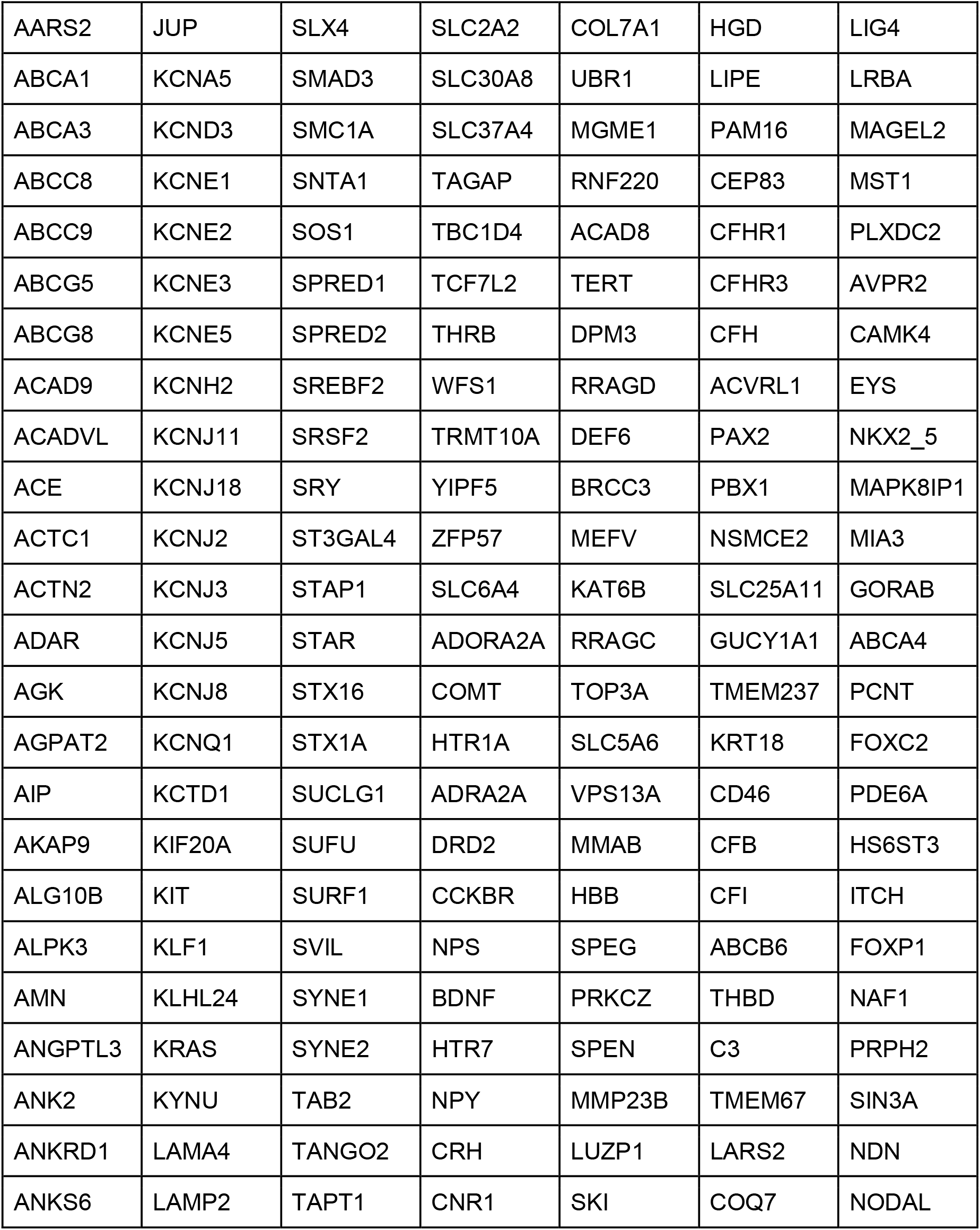
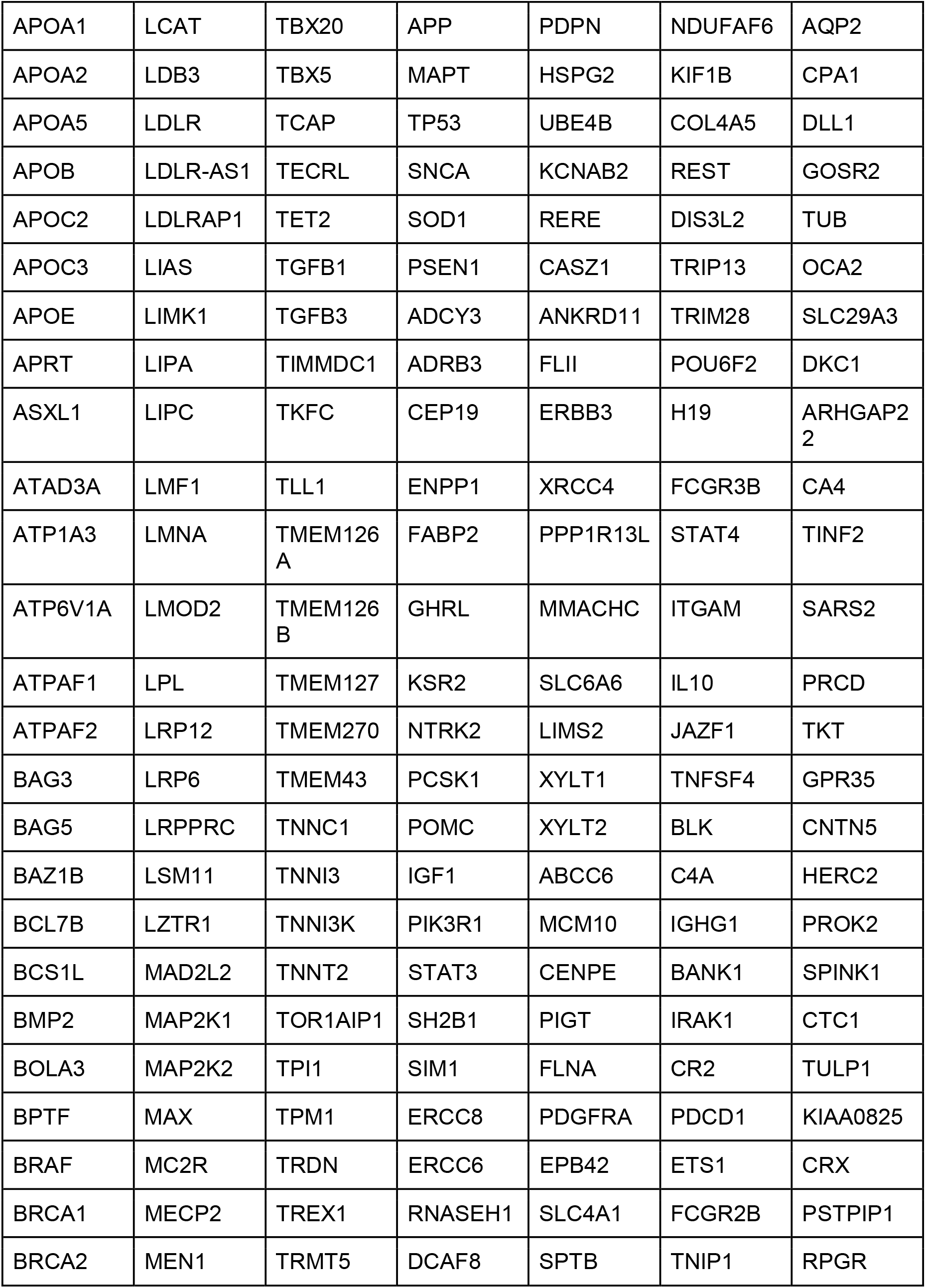
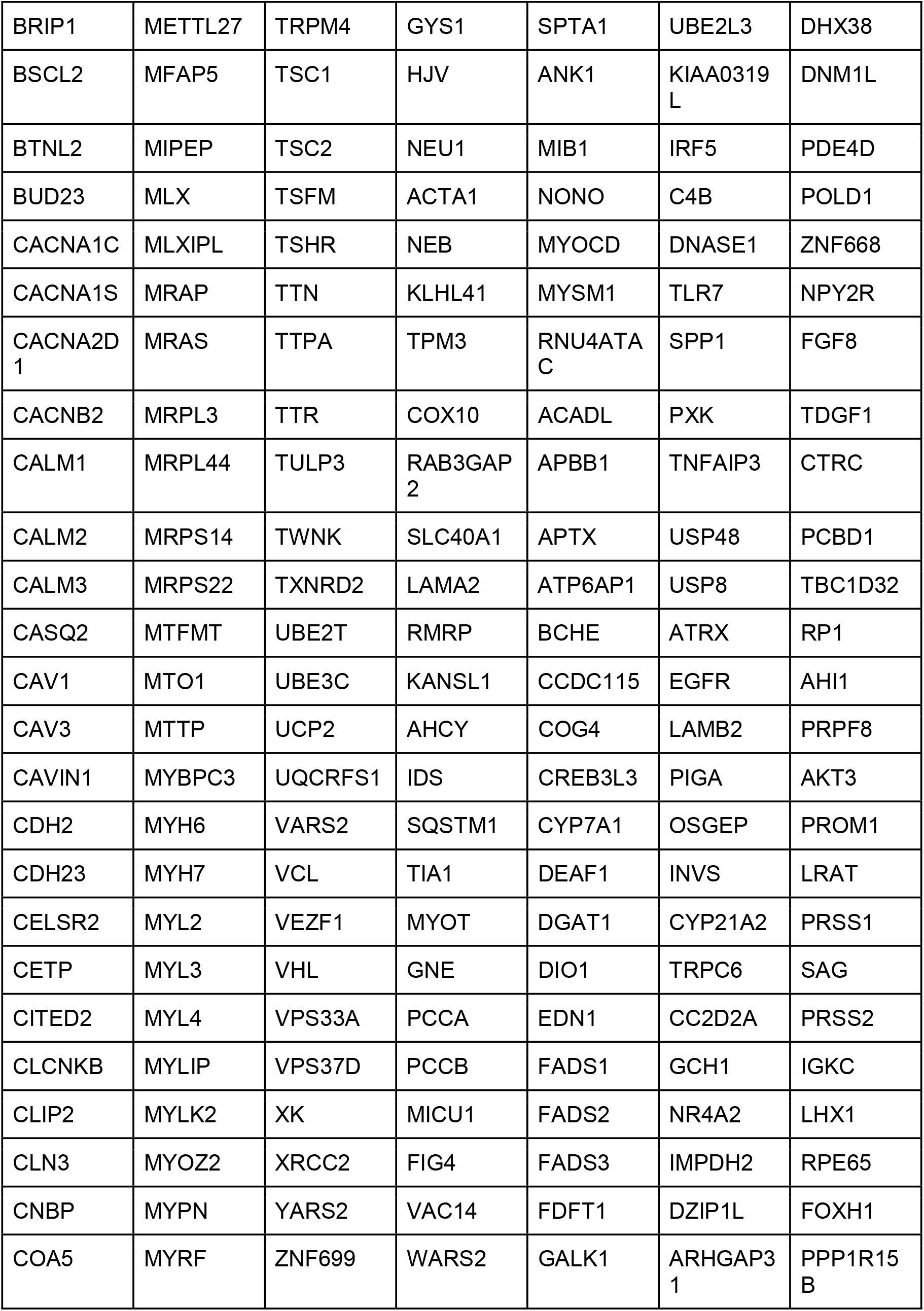
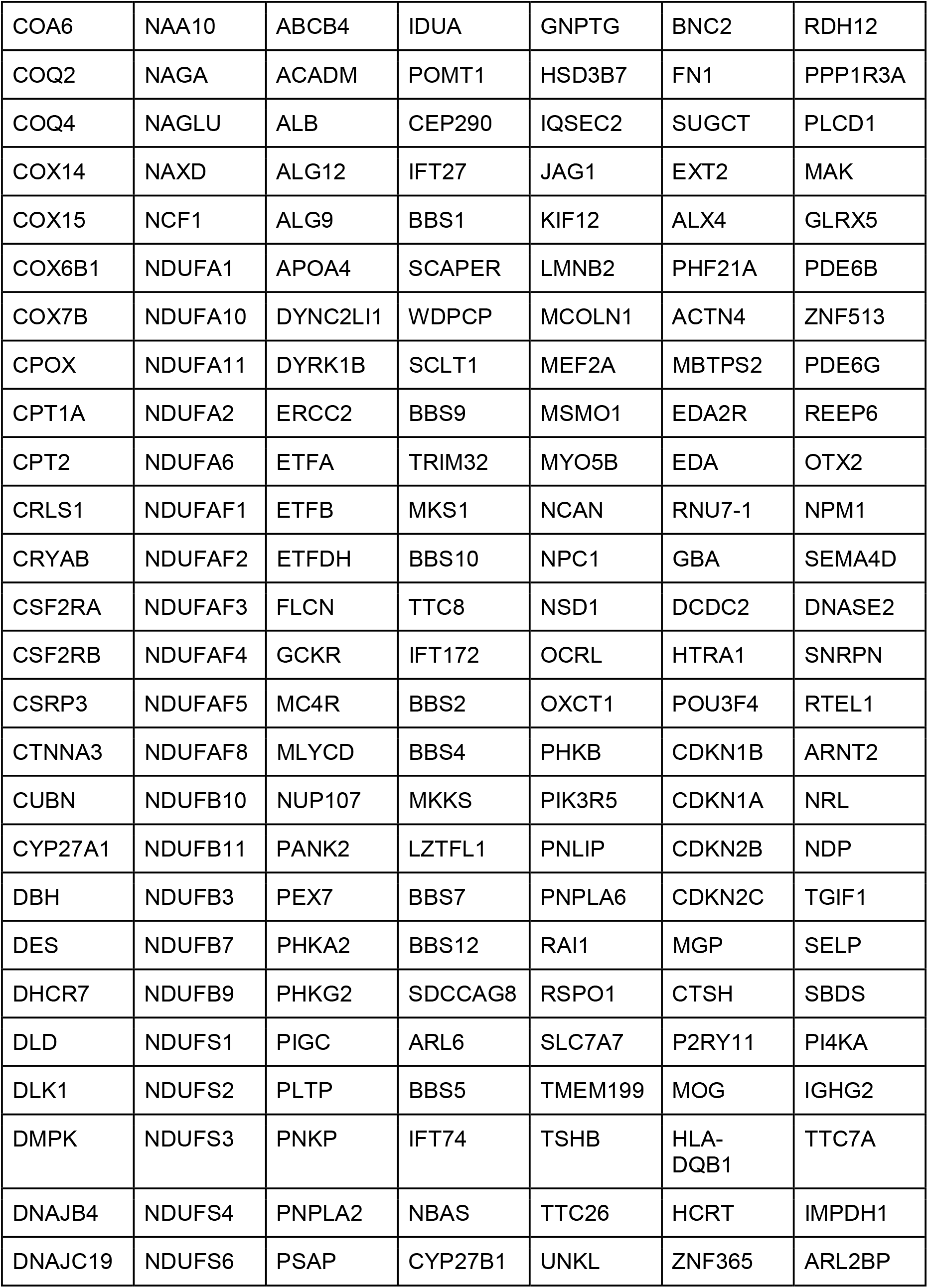
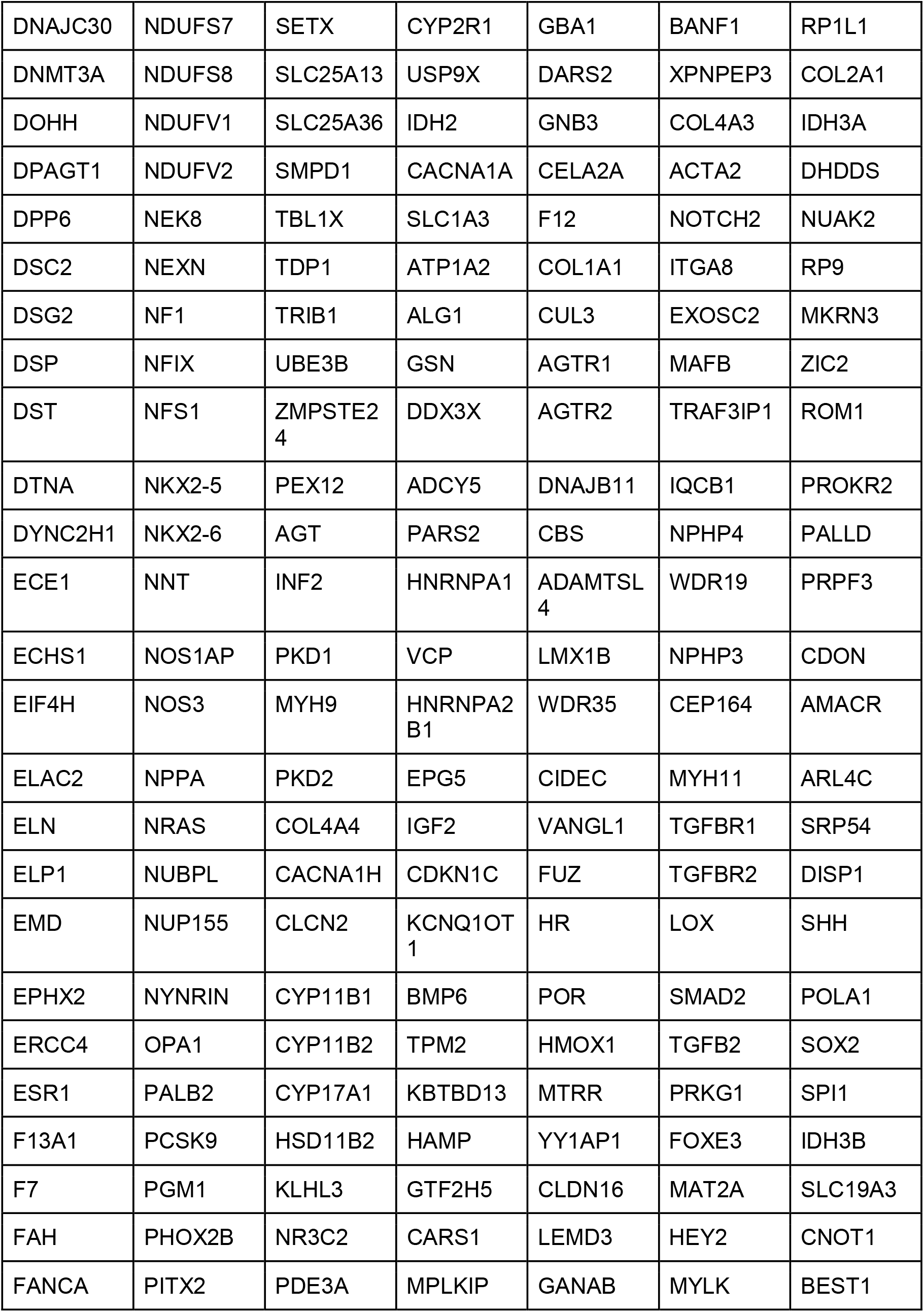
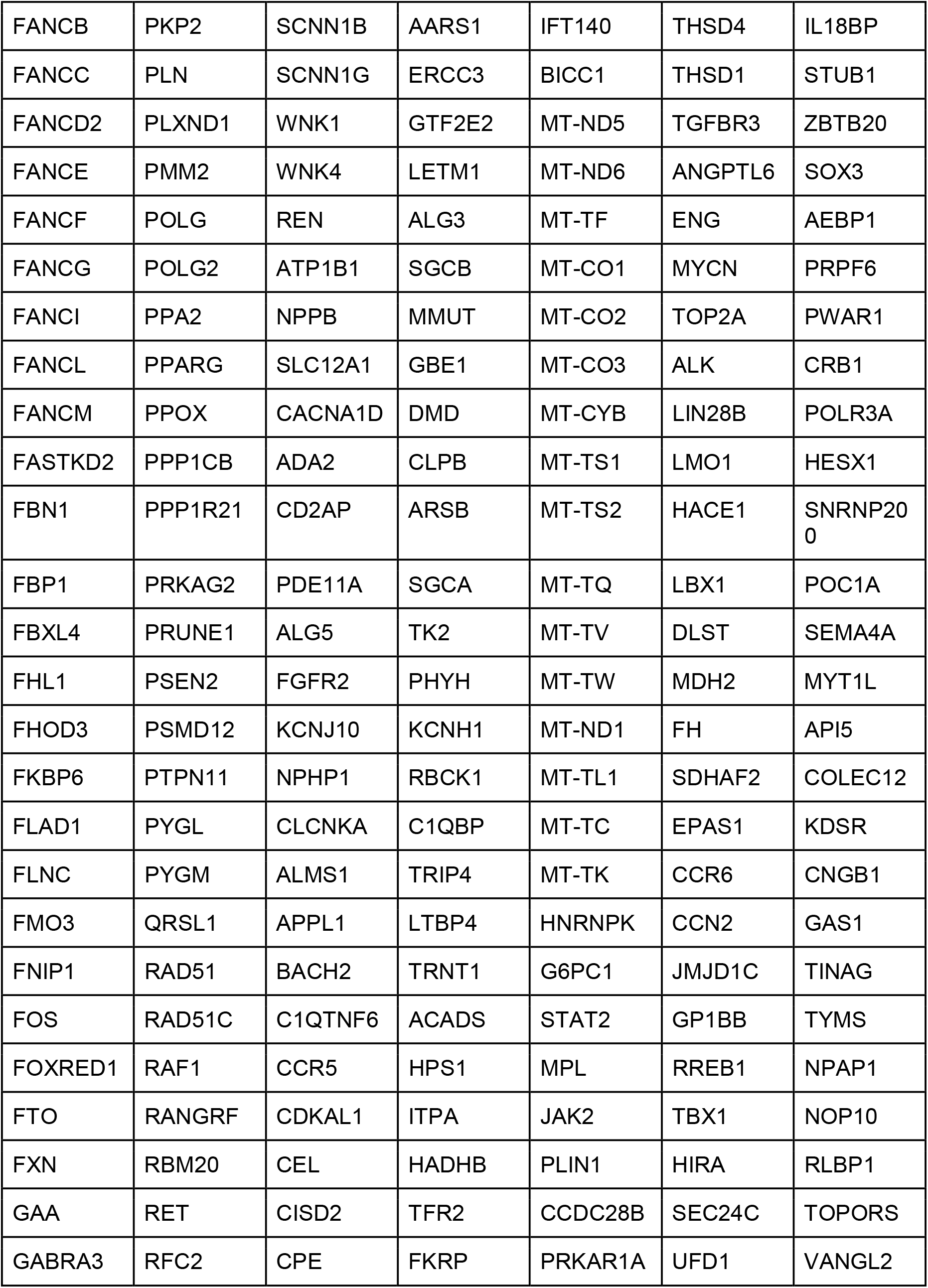
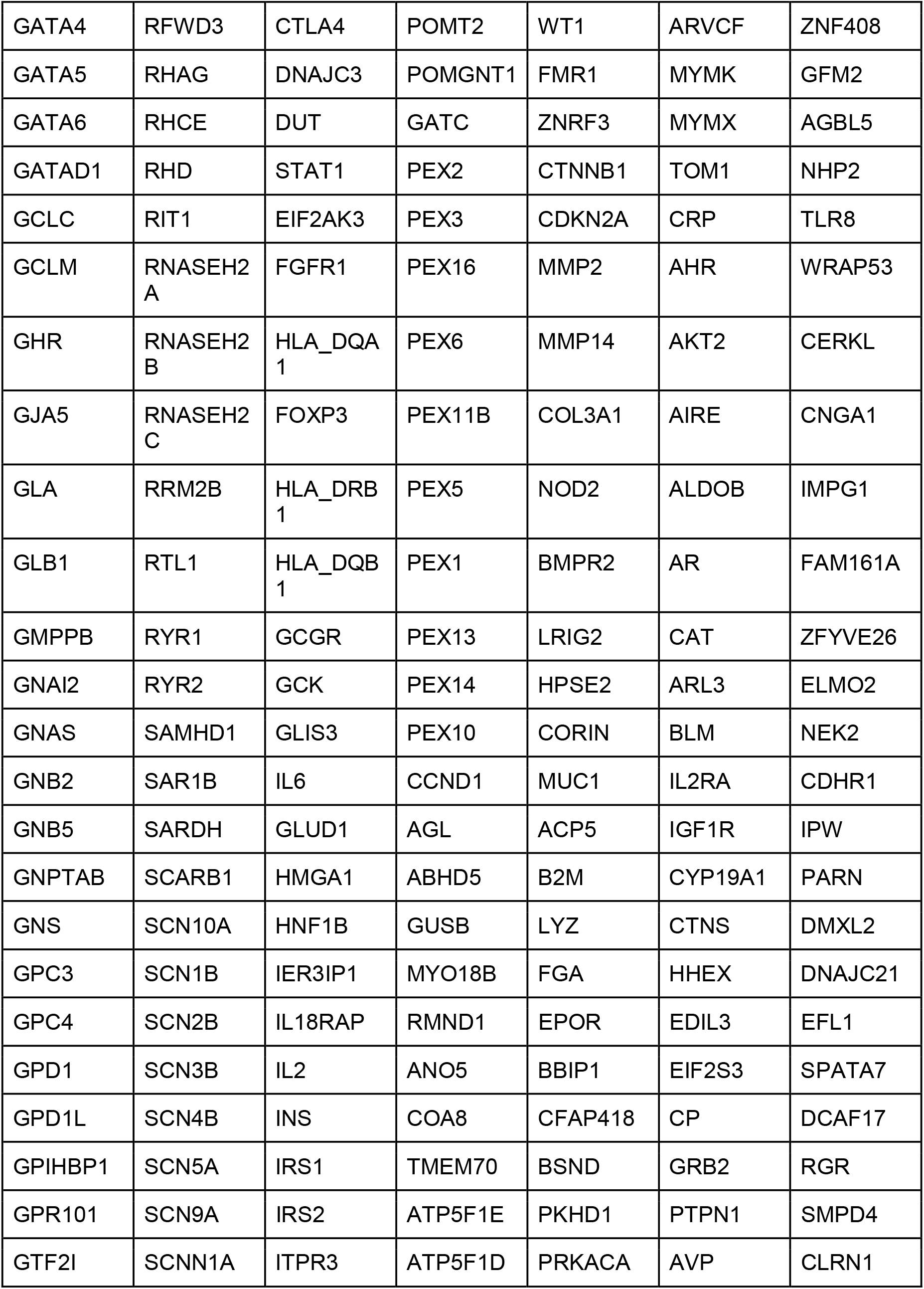
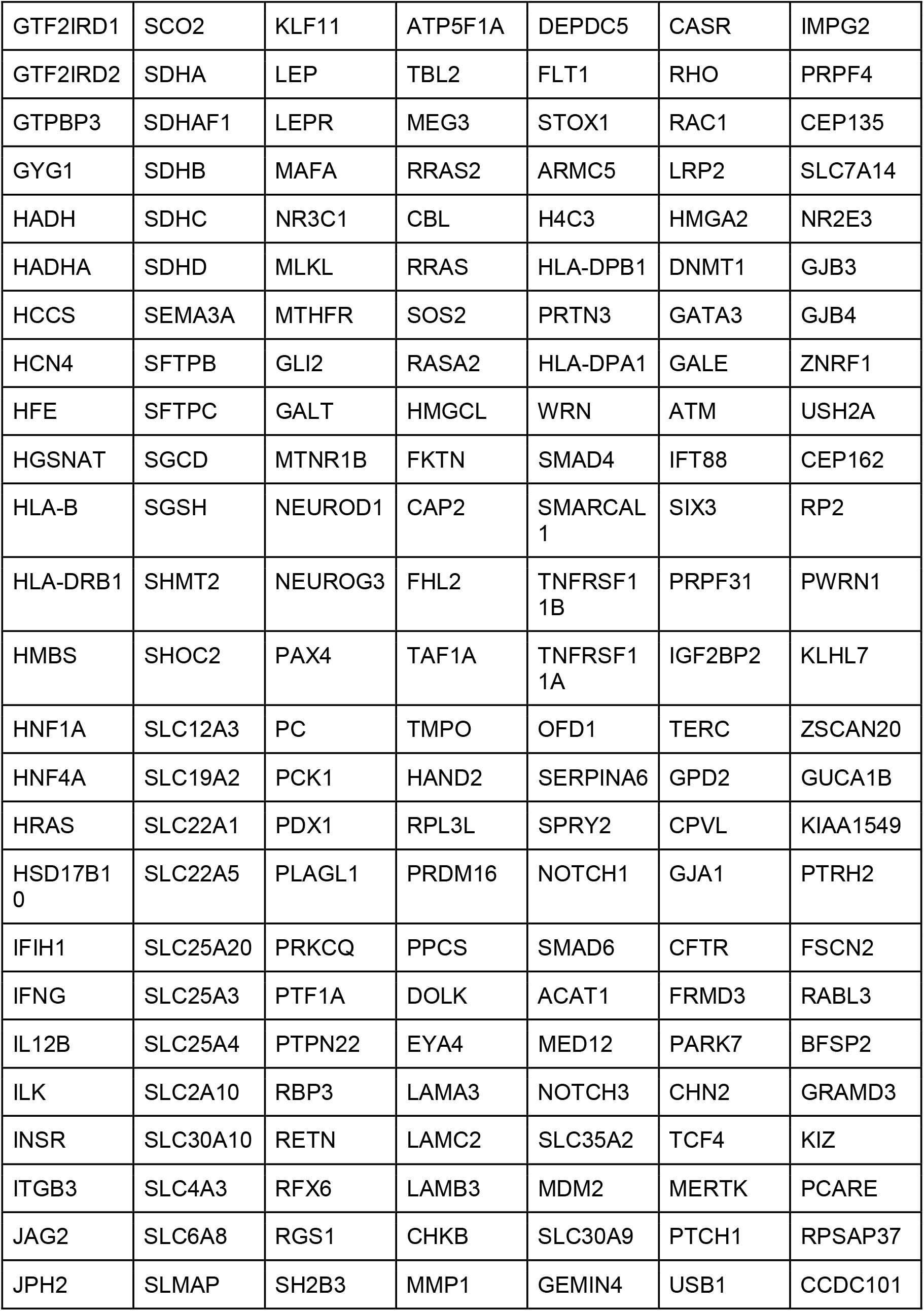
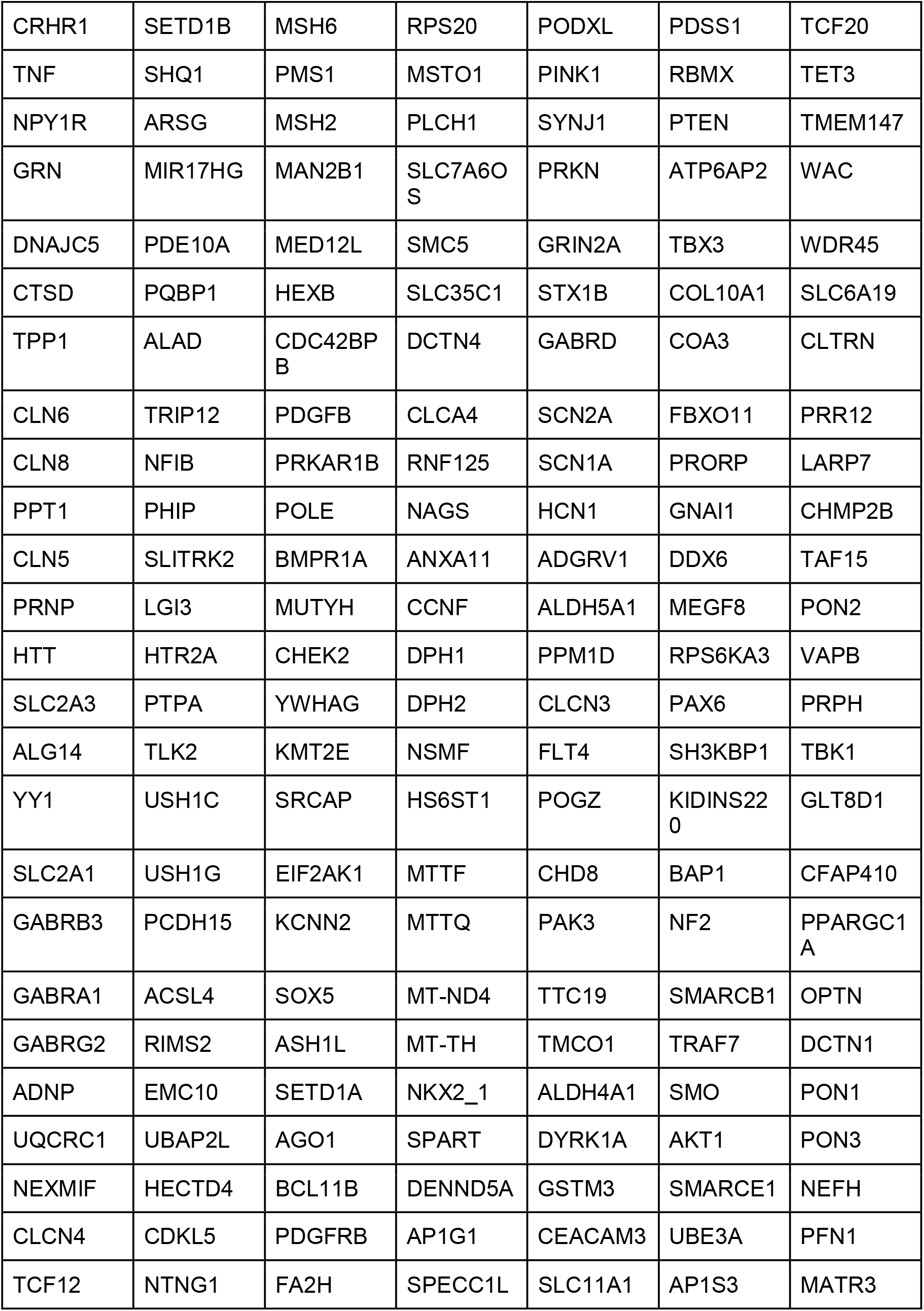
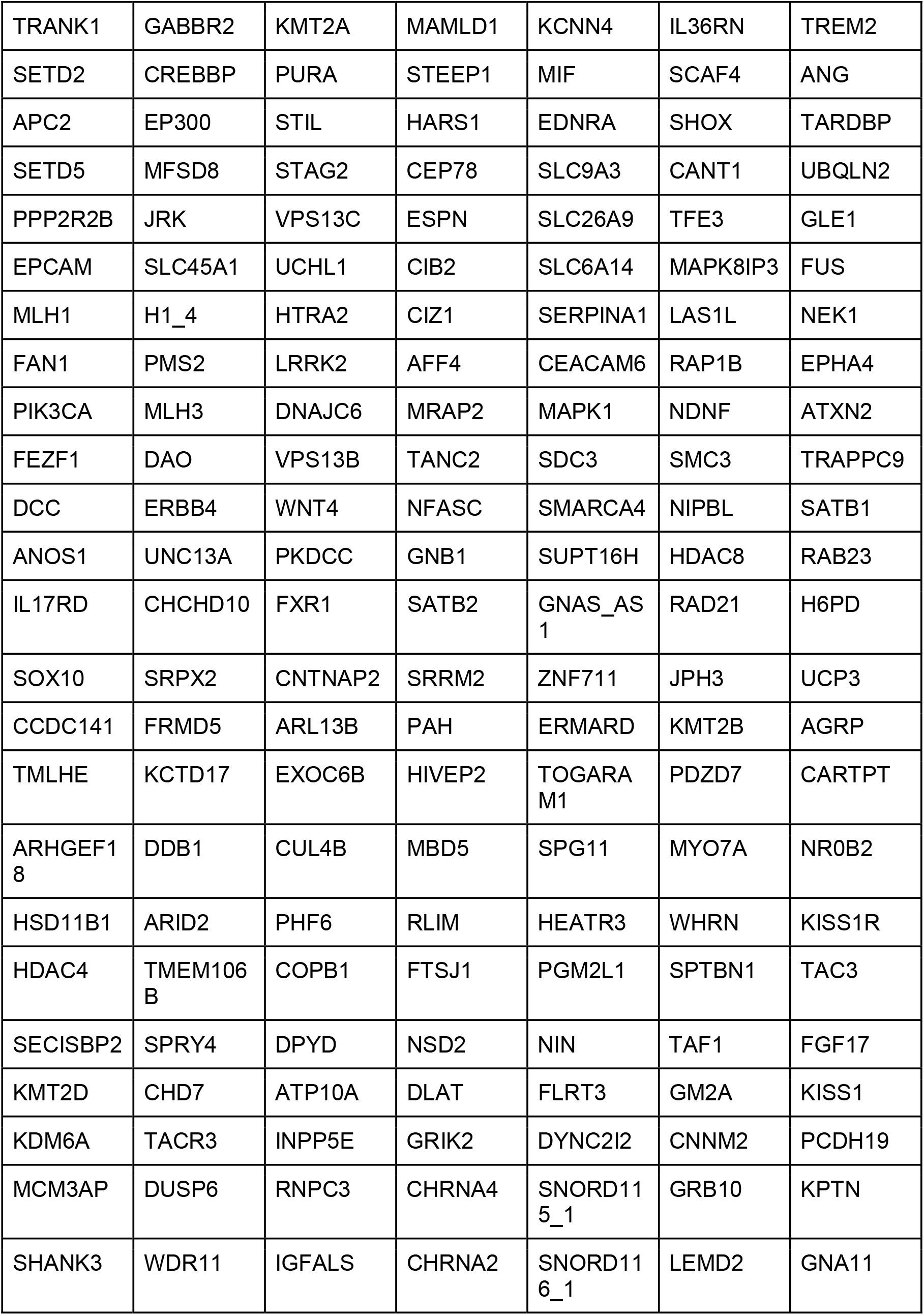
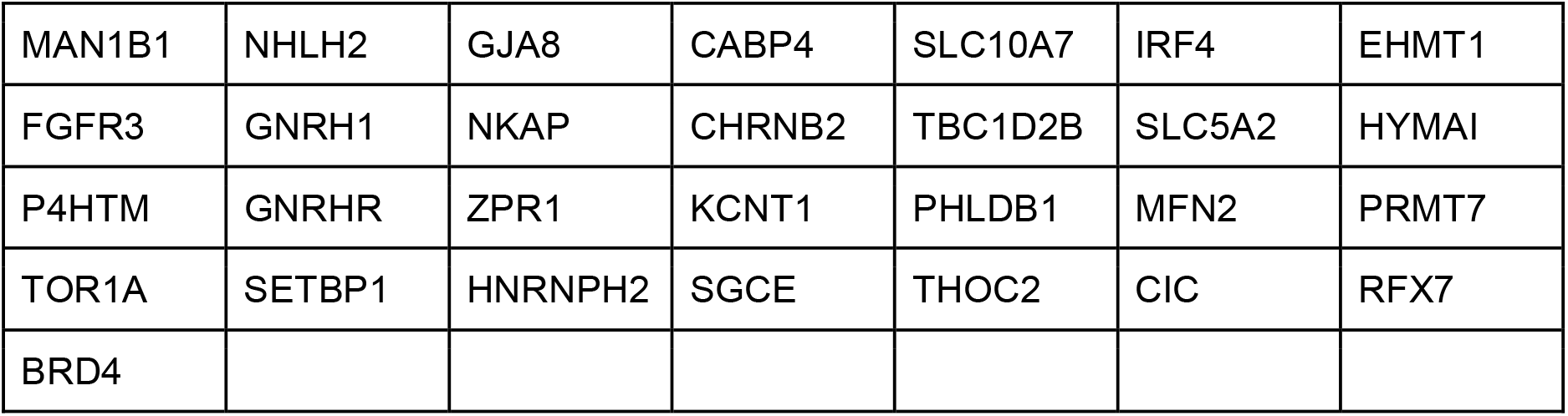
CAD Associated Genes.

Values are presented as mean ± approximate SD, with corresponding 95% CIs. The low SDs and narrow CIs reflect stable and consistent model behavior on the training data given in Table 4 and testing data given in Table 3.

#### Robustness and Generalizability Assessment

To assess the model’s reliability, we performed **200 bootstrap iterations** on the final integrated model. For each iteration, key classification metrics were evaluated across both training and test sets.

As shown in **Figure 8**, each point represents the mean of a metric, while the **horizontal error bars depict 95% confidence intervals (CIs)**.

**Figure 8:**
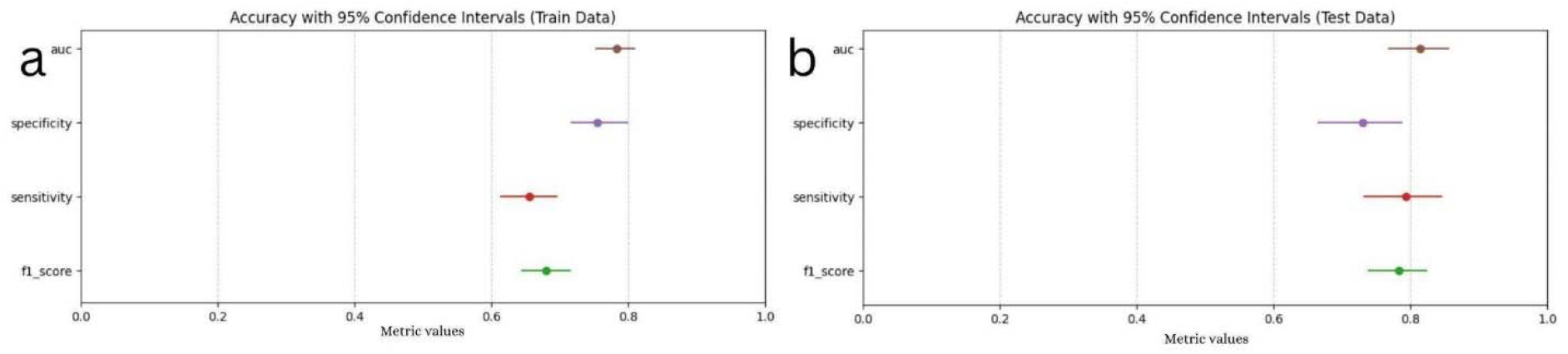
Performance metrics with 95 % Confidence Intervals. (a) Training data and (b) test data results show consistent performance across key metrics — AUC, specificity, sensitivity, and F1 score — with tight confidence intervals. The model demonstrates robust and reliable classification performance across both train and test datasets.

- The **narrow confidence intervals** and **low standard deviations** observed across all metrics indicate a **high degree of model stability** and minimal performance fluctuation due to random variation.

This strongly supports the model’s **generalizability** to unseen data and affirms its reproducibility.

#### Error Analysis

An analysis of misclassified samples in the final stacked model revealed:

- **False Positives:** 12.5%
- **False Negatives:** 11.2%

This relatively balanced error distribution suggests the model does not overfit to one class, making it reliable for both community screening and clinical prioritization use cases. Direct PRS computation was not performed in this cohort. Instead, benchmarking is framed as a future direction, referencing canonical PRS literature (e.g., Khera et al., Mars et al.). This is transparently reported to prevent overstatement of model scope.

## Discussion

Coronary artery disease (CAD) remains a major cause of mortality worldwide, especially in low- and middle-income countries like India, where early onset and higher morbidity rates amplify its socio-economic burden [7, 19]. Traditional risk prediction models—largely developed in Western cohorts— focus predominantly on modifiable (e.g., lifestyle, smoking, diet) and non-modifiable (e.g., age, gender, family history) factors, along with biochemical markers such as lipid profiles and HbA1c [8, 20]. However, these models inadequately incorporate population-specific genetic and epigenetic contributions, especially in South Asian populations where CAD often presents at younger ages and with greater severity [21].

The heritable component of CAD is now well-recognized, with studies suggesting that genetics can explain 40–60% of the disease variance [9, 22]. Polygenic risk scores (PRS) derived from genome-wide association studies (GWAS) offer a promising route for personalized risk assessment [15, 23]. However, most PRS models are built using datasets predominantly from European ancestry, which limits their clinical translation to ethnically diverse populations [24].

Our study aims to bridge this gap by introducing a robust, AI-integrated framework that tailors genetic risk prediction to the South Indian population using whole exome sequencing (WES). We employed **GEMS**, an LLM-based literature mining tool, to score 1772 genes based on their association strength with CAD, epigenetic regulation, and diabetes-associated macrovascular complications. The scoring system (1–9) enables prioritization of genes for downstream variant analysis.

To enhance variant interpretation, we developed **CASCADE**, a tiered consequence assessment model that integrates variant type (common, rare, novel), copy number variations (CNVs), and population-specific minor allele frequencies (MAFs). Since most MAFs are not readily available for the Indian population, we derived them from our cohort of 1,243 individuals. This aspect addresses the challenge of allele frequency misrepresentation, a key limitation in applying global PRS to non-European populations [14, 25].

Further, the **SIGMA** algorithm—our machine learning-based risk stratification model—demonstrates that CAD risk prediction improves significantly when genomic data is contextualized with demographic parameters such as age and gender. This aligns with the clinical necessity of applying prevention strategies at age-appropriate milestones [18, 26]. For example, while a 24-year-old female may genetically fall in a higher-risk bracket, the actionable clinical recommendation should be time-bound— e.g., cardiology follow-up beginning at age 50—thereby improving the relevance of intervention.

Additionally, the inclusion of epigenetic regulation—while currently limited due to the use of WES—is a forward-thinking aspect of this study. Epigenetic modulators such as non-coding RNAs, methylation patterns, and histone modifications influence gene expression and phenotypic presentation in CAD but are missed in WES datasets [5, 13, 27]. Expanding this research into whole genome sequencing (WGS) and methylome profiling would allow for more accurate modeling, especially in early-onset or idiopathic cases.

Another key innovation lies in SIGMA’s modular architecture, which allows population-specific fine-tuning. Similar frameworks have demonstrated success in other diseases such as asthma, COPD, and diabetes [2, 5, 28], indicating the scalability of our approach. Our model’s performance—**79% sensitivity and 72% specificity** for CAD risk prediction—shows promising translational value for community screening and precision prevention in South India.

Nonetheless, this study has limitations. First, **WES lacks coverage of non-coding regulatory regions**, limiting detection of enhancers, promoters, and long non-coding RNAs (lncRNAs) which play critical roles in gene expression and disease manifestation [13, 29]. Second, although our sample size is adequate for model training, external validation in larger, independent cohorts is necessary. Third, the **clinical relevance of rare vs. common variants** remains debated, and more functional annotation studies are required to clarify their roles [16, 30]. Finally, integrating traditional, lifestyle, and biochemical parameters into SIGMA could significantly enhance prediction accuracy—an area for future enhancement.

## Conclusions

This study is the first attempt to bring in AI/ML models in accurate risk stratifying CAD risk for a given individual using whole exome data. We developed a model of scoring 1772 genes using GEMS, weighing in all variants in these genes using an integrated tool called CASCADE, and developing ML algorithm SIGMA to risk stratify each individual into low, mild, moderate, high categories for developing CAD with 79% sensitivity (SD ±0.0292) and 72% (SD ±0.0313) specificity for south Indian Population.

### Limitations

- Whole genome sequencing data is ideal to perform in population studies but because of cost efficiency, Whole exome sequencing was performed.
- The Clinical importance of common vs rare variants is still debatable.
- A Sample size of 1243 is good for model development
- Epigenetic regulators at the genome level are mostly noncoding areas, RNAs, and Histone complexes which are not covered in Exomes.
- More accurate predictions can be made if we can incorporate all traditional and biochemical risk factors into SIGMA.
- Age imbalance between cases and controls: CAD cases were significantly older than controls. Although age was modeled explicitly and sensitivity analyses were performed, residual confounding remains possible.
- Incomplete availability of clinical risk factors Traditional variables such as smoking, diabetes, hypertension, lipid profile, and BMI were inconsistently recorded and therefore excluded from the model. Their absence likely reduced predictive performance and should be integrated in future work.
- Lack of direct PRS benchmarking: Polygenic risk scores were not computed for this dataset. Direct comparison with established PRS frameworks would provide complementary insight but is planned for future research.
- Limited environmental and lifestyle variables CAD risk reflects both genetic and non-genetic factors. The lack of detailed environmental and behavioral data (nutrition, exercise, socioeconomic metrics) constrains interpretation of gene–environment interaction effects.

Authors have no competing interests.

## Funding

None

## Data and Code availability

De-identified genomic and annotation files generated during this study — including VCF, CSV, and JSON formats — are available upon reasonable request to the corresponding author. The curated list of 1772 CAD-associated genes is provided as Supplementary Table 5.

Due to proprietary constraints, the full GEMS and CASCADE pipelines cannot be shared, but high-level methods are described in sufficient detail to enable conceptual replication.

## Ethics approval and consent to participate

All participants provided **written informed consent** permitting the use of de-identified genomic and phenotypic data for research and development. Data were originally generated as part of routine clinical genetic testing. No institutional review board (IRB) approval was applicable because all data were fully anonymized prior to analysis, in accordance with regional regulatory guidelines for de-identified clinical datasets.

Participants were informed that **genomic data may be used for research**, and a standard clinical consent template is available upon request.

## Consent for publication

Yes

## Authors’ contributions

K.K.V. (Krishna Kumar Vempati) and L.N.K. (Lakshmi Nirmala Kadali): Designed and implemented the methodology, conducted data analysis, and led the machine learning model development and evaluation. K.R.U. (Kalyan Ram Uppaluri) and H.J.C. (Hima Jyothi Challa): Conceived the study concept, contributed to manuscript drafting, and refined the scientific narrative.V.B. (Venu Battena) and A.K. (Alekhya Kosuri): Provided bioinformatics support, including data preprocessing, variant annotation, and workflow integration.

## Acknowledgements

None

## Nomenclature

SD: Standard deviation
CI: Confidence interval
AUC: Area under the curve
ROC: Receiver Operating Characteristic
CAD: Coronary Artery Disease
CHROM: Chromosome
POS: Position
REF: Reference
ALT: Alterative
rsID: Variant reference ID
WES: Whole Exome Sequencing
PCI: percutaneous coronary intervention
GENEINFO: Gene information
AF: Allele Fraction
ML: Machine Learning
BED: 
ACMG: American College of Medical Genetics and Genomics
GEMS: GeneConnectRx Evidence Metrics
NCBI: National Center for Biotechnology Information
CASCADE: Comprehensive Assessment of Sequence and Clinical Annotation Data Evaluation
SIGMA: Scoring Importance of Gene Specific to Disease applying Advanced ML algorithms
TP: True positive
FP: False positive
TN: True Negative
FN: False Negative
VCF: Variant Call format
MAF: Minor Allele Frequency
VAF: Variant Allele Frequency
CNV: Copy Number Variation
SIFT: Sorting Intolerant From Tolerant
PolyPhen: Polymorphism Phenotyping
VEP: Variant Effect Predictor
DP: Depth
VAF: Variant allele fraction
LLM: Large Language Model
OR: Odds Ratio

## Notes

### Competing Interest Statement

The authors have declared no competing interest.

### Funding Statement

This study did not receive any funding

### Author Declarations

Ethics Committee of Uppaluri K&H Personalised Medicine Clinic Hyderabad waived ethical approval for this work, as the study used only patient-consented, fully de-identified data, and the SIGMA-ML platform was developed and validated exclusively on anonymized datasets with no access to identifiable patient information.

## References

1. Wellcome Trust Case Control Consortium. Genome-wide association study of 14,000 cases of seven common diseases and 3,000 shared controls. Nature. 2007 Jun 7;447(7145):661–78. doi: 10.1038/nature05911. PMID: 17554300; PMCID: PMC2719288.

2. Moll M, Sakornsakolpat P, Shrine N, Hobbs BD, DeMeo DL, John C, Guyatt AL, McGeachie MJ, Gharib SA, Obeidat M, Lahousse L, Wijnant SRA, Brusselle G, Meyers DA, Bleecker ER, Li X, Tal-Singer R, Manichaikul A, Rich SS, Won S, Kim WJ, Do AR, Washko GR, Barr RG, Psaty BM, Bartz TM, Hansel NN, Barnes K, Hokanson JE, Crapo JD, Lynch D, Bakke P, Gulsvik A, Hall IP, Wain L; International COPD Genetics Consortium; SpiroMeta Consortium; Weiss ST, Silverman EK, Dudbridge F, Tobin MD, Cho MH. Chronic obstructive pulmonary disease and related phenotypes: polygenic risk scores in population-based and case-control cohorts. Lancet Respir Med. 2020 Jul;8(7):696–708. doi: 10.1016/S2213-2600(20)30101-6. Erratum in: Lancet Respir Med. 2024 Nov;12(11):e70. doi: 10.1016/S2213-2600(24)00326-6. PMID: 32649918; PMCID: PMC7429152.

3. Goldstein DB. Common genetic variation and human traits. N Engl J Med. 2009 Apr 23;360(17):1696–8. doi: 10.1056/NEJMp0806284. Epub 2009 Apr 15. PMID: 19369660.

4. Hirschhorn JN. Genomewide association studies--illuminating biologic pathways. N Engl J Med. 2009 Apr 23;360(17):1699–701. doi: 10.1056/NEJMp0808934. Epub 2009 Apr 15. PMID: 19369661.

5. Forno E, Wang T, Qi C, Yan Q, Xu CJ, Boutaoui N, Han YY, Weeks DE, Jiang Y, Rosser F, Vonk JM, Brouwer S, Acosta-Perez E, Colón-Semidey A, Alvarez M, Canino G, Koppelman GH, Chen W, Celedón JC. DNA methylation in nasal epithelium, atopy, and atopic asthma in children: a genome-wide study. Lancet Respir Med. 2019 Apr;7(4):336–346. doi: 10.1016/S2213-2600(18)30466-1. Epub 2018 Dec 21. PMID: 30584054; PMCID: PMC6441380.

6. https://www.ncbi.nlm.nih.gov/research/pubtator3/

7. Sharma M, Ganguly NK. Premature coronary artery disease in Indians and its associated risk factors. Vasc Health Risk Manag. 2005;1(3):217-25. PMID: 17319107; PMCID: PMC1993956.

8. Hajar R. Risk Factors for Coronary Artery Disease: Historical Perspectives. Heart Views. 2017 Jul-Sep;18(3):109–114. doi: 10.4103/HEARTVIEWS.HEARTVIEWS_106_17. PMID: 29184622; PMCID: PMC5686931.

9. Roberts R, Chang CC, Hadley T. Genetic Risk Stratification: A Paradigm Shift in Prevention of Coronary Artery Disease. JACC Basic Transl Sci. 2021 Mar 22;6(3):287–304. doi: 10.1016/j.jacbts.2020.09.004. PMID: 33778213; PMCID: PMC7987546.

10. Hyunok Yun, Nan Iee Noh, Eun Young Lee. Genetic risk scores used in cardiovascular disease prediction models: a systematic review. Rev. Cardiovasc. Med. 2022, 23(1), 8. 10.31083/j.rcm2301008.

11. Muse ED, Chen SF, Torkamani A. Monogenic and Polygenic Models of Coronary Artery Disease. Curr Cardiol Rep. 2021 Jul 1;23(8):107. doi: 10.1007/s11886-021-01540-0. PMID: 34196841; PMCID: PMC8317496.

12. Dai X, Wiernek S, Evans JP, Runge MS. Genetics of coronary artery disease and myocardial infarction. World J Cardiol. 2016 Jan 26;8(1):1–23. doi: 10.4330/wjc.v8.i1.1. PMID: 26839654; PMCID: PMC4728103.

13. Shi Y, Zhang H, Huang S, Yin L, Wang F, Luo P, Huang H. Epigenetic regulation in cardiovascular disease: mechanisms and advances in clinical trials. Signal Transduct Target Ther. 2022 Jun 25;7(1):200. doi: 10.1038/s41392-022-01055-2. PMID: 35752619; PMCID: PMC9233709.

14. Kido, T., Sikora-Wohlfeld, W., Kawashima, M. et al. Are minor alleles more likely to be risk alleles?. BMC Med Genomics 11, 3 (2018). 10.1186/s12920-018-0322-5.

15. Duncan L, Shen H, Gelaye B, Meijsen J, Ressler K, Feldman M, Peterson R, Domingue B. Analysis of polygenic risk score usage and performance in diverse human populations. Nat Commun. 2019 Jul 25;10(1):3328. doi: 10.1038/s41467-019-11112-0. PMID: 31346163; PMCID: PMC6658471.

16. Gibson G. Rare and common variants: twenty arguments. Nat Rev Genet. 2012 Jan 18;13(2):135–45. doi: 10.1038/nrg3118. PMID: 22251874; PMCID: PMC4408201.

17. Shriver MD, Smith MW, Jin L, Marcini A, Akey JM, Deka R, Ferrell RE. Ethnic-affiliation estimation by use of population-specific DNA markers. Am J Hum Genet. 1997 Apr;60(4):957-64. PMID: 9106543; PMCID: PMC1712479.

18. Hung DY, Rundall TG, Tallia AF, Cohen DJ, Halpin HA, Crabtree BF. Rethinking prevention in primary care: applying the chronic care model to address health risk behaviors. Milbank Q. 2007;85(1):69–91. doi: 10.1111/j.1468-0009.2007.00477.x. PMID: 17319807; PMCID: PMC2690311.

19. Prabhakaran D, Jeemon P, Roy A. Cardiovascular Diseases in India. Circulation. 2016 Apr 19;133(16):1605–20. doi:10.1161/CIRCULATIONAHA.114.008729

20. Yusuf S, Hawken S, Ounpuu S, et al. Effect of potentially modifiable risk factors associated with myocardial infarction in 52 countries (the INTERHEART study): case-control study. Lancet. 2004;364(9438):937–52.

21. Joshi P, Islam S, Pais P, et al. Risk factors for early myocardial infarction in South Asians compared with individuals in other countries. JAMA. 2007;297(3):286–94.

22. Schunkert H, et al. Large-scale association analysis identifies 13 new susceptibility loci for coronary artery disease. Nat Genet. 2011 Mar;43(4):333–8.

23. Khera AV, et al. Genome-wide polygenic scores for common diseases identify individuals with risk equivalent to monogenic mutations. Nat Genet. 2018 Sep;50(9):1219–24.

24. Martin AR, et al. Clinical use of current polygenic risk scores may exacerbate health disparities. Nat Genet. 2019;51(4):584–91.

25. Sirugo G, Williams SM, Tishkoff SA. The missing diversity in human genetic studies. Cell. 2019 Mar 21;177(1):26–31.

26. Lloyd-Jones DM, et al. Lifetime risk prediction and the implications for prevention. Circ Cardiovasc Qual Outcomes. 2010;3(6):626–34.

27. Greco CM, Condorelli G. Epigenetic modifications and noncoding RNAs in cardiac hypertrophy and failure. Nat Rev Cardiol. 2015 Aug;12(8):488–97.

28. Ober C, Vercelli D. Gene-environment interactions in human disease: nuisance or opportunity? Trends Genet. 2011 Mar;27(3):107–15.

29. Maurano MT, et al. Systematic localization of common disease-associated variation in regulatory DNA. Science. 2012 Sep 7;337(6099):1190–5.

30. Manolio TA, et al. Finding the missing heritability of complex diseases. Nature. 2009 Oct 8;461(7265):747–53.

